# A Linear Analysis Mixed Model Phenotyper (LAMP) for Bipolar Disorder: Evaluating the Impact of Medication, Substance Use Disorders, and Comorbidities on Mood Trajectories

**DOI:** 10.1101/2025.09.06.25335235

**Authors:** Qingzhi Liu, Pranjal Srivastava, Ivan Rozhkov, Gregory A. Farnum, Monica J. Holmes, Lars G. Fritsche, David Belmonte, Alex Ade, Jie Sun, Anastasia Yocum, Melvin Mclnnis, Veera Baladandayuthapani, Brian D. Athey

## Abstract

**Background:** Bipolar disorder is a complex psychiatric condition characterized by recurrent mood episodes and fluctuating symptom severity. Our study investigates post-diagnosis complexities using 17-year longitudinal data from the Prechter Bipolar Research Program.

**Methods:** We analyzed mood trajectories of 525 bipolar I disorder (BP1) and 150 bipolar II disorder (BP2) patients, using the Linear Analysis Mixed Model Phenotyper (LAMP) to identify subgroups. This approach revealed shared and distinct patterns across individual trajectories, classifying these subgroups into meaningful longitudinal phenotypes. Regression modeling was then applied to identify associations between these phenotypes and relevant factors.

**Results:** We found that medication use and alcohol use disorder significantly impacted long-term mood trajectories. Polypharmacy was associated with resistant/worsening trajectories in both BP1 (PHQ-9: OR: 1.09, CI: 1.02–1.17) and BP2 (GAD-7: OR: 1.21, CI: 1.05–1.43), with each additional medication increasing the risk of a worse trajectory by 9% and 21%, respectively. In BP1, antidepressant use improved trajectories (PHQ-9: OR: 0.40, CI: 0.19-0.83), but when combined with antipsychotics, increasing use frequency from 0% to 20% raised the risk of resistant/worsening trajectories (OR: 2.09, CI: 1.33– 3.28). In BP2, antipsychotic use was associated with improved trajectories (PHQ-9: OR: 0.04, CI: 0.00-0.56). Alcohol use disorder in BP1 increased resistant/worsening risk by 83% (PHQ-9: OR: 1.83, CI: 1.15-2.93).

**Conclusions:** Our results indicate that polypharmacy and alcohol use disorder contribute to resistant or worsening mood trajectories. In BP1, while antidepressants are beneficial, their combination with antipsychotics substantially increases risk, highlighting the need for individualized, diagnosis-specific treatment approaches.

**Highlights:**

- LAMP analysis pipeline aided in improving treatment approaches in bipolar disorder.
- 17 years of PrBP data provided comprehensive analysis of long-term mood trajectories.
- Polypharmacy and alcohol use disorder worsened mood trajectory.
- The combination of antidepressants and antipsychotics worsened mood trajectory.

## 1. Introduction

Bipolar disorder (BD) represents a significant health challenge, affecting over 1% of the global population (Merikangas et al., 2011). It is a chronic psychiatric condition characterized by recurrent episodes of mania, which include elevated mood, hyperactivity, and sometimes psychosis, and episodes of depression. BD often leads to substantial functional impairment and a reduced quality of life as patients typically experience a wide range of challenging clinical outcomes and mood trajectories (Oldis et al., 2016).

BD is classified into two types: Bipolar I disorder (BP1), characterized by at least one episode of mania, often with psychotic symptoms, and major depressive episodes; and Bipolar II disorder (BP2), which involves recurrent major depressive episodes and hypomania, a milder form of mania without psychotic symptoms and less severe disruption to daily life (Vieta et al., 2018). The complex etiology, variable clinical presentations, and unpredictable course of BD complicate its management (Mignogna and Goes, 2024). Furthermore, varied medication regimens and common comorbidities, such as anxiety and substance use disorders, exacerbate these challenges. Approximately 45% of individuals with BD have comorbid anxiety disorders, and substance use disorders are prevalent, leading to poorer outcomes and higher suicide risk (Merikangas et al., 2007; Vieta et al., 2018).

BD is lifelong and highly heterogeneous at the person level. In 13-year follow-ups, patients with BP1 and BP2 are symptomatic during 47.3% and 53.9% of weeks, respectively (Judd et al., 2003, 2002). Additionally, clinical courses also diverge markedly: some patients improve, whereas others show persistent or worsening symptoms (Mignogna and Goes, 2024). This variability, both between persons and within persons, together with polypharmacy, substance use disorders, and medical and psychiatric comorbidities makes it difficult to determine which factors truly shape mood score trajectories and individual treatment response.

Longitudinal cohort studies have begun to link patient characteristics with course. In STEP-BD, poorer trajectories across depression, mania, and functioning are associated with lower education, unemployment, and greater comorbidity (Mignogna and Goes, 2024). In youth, COBY shows that later symptom onset, less extensive family histories of BD and substance use disorders, and milder baseline symptoms predict more favorable trajectories of euthymia and cognitive functioning (Birmaher et al., 2014; Frías et al., 2017). Similarly, Weintraub et al. (2020) found that youth belonging to minority races and those with more severe baseline depressive symptoms, suicidal ideation, and lower quality of life scores experienced more symptomatic courses of illness over time. These findings support the need for individualized care, but gaps remain. Few studies extend beyond ten years, and the long-term effects of patient factors and medication exposure on mood trajectories are not well defined.

The Heinz C. Prechter Bipolar Research Project (PrBP), with its 17-year duration and bi-monthly mood assessments, provided a uniquely detailed dataset compared to other large bipolar disorder (BD) cohorts (Yocum et al., 2023). Its rich data, including mood scores, annual medication records, and diverse phenotypes such as demographics, substance use disorders, and comorbidities, allowed for precise examination of the complex, long-term dynamics of BD. Prior analyses of PrBP data have leveraged longitudinal mood scores to examine associations with suicidal ideation (Smith et al., 2025), the interplay between anxiety and depression (Kim et al., 2024), and alcohol use and functioning (Sperry et al., 2024). While prior PrBP studies have provided valuable insights, the long-term influence of diverse clinical and demographic features on the trajectories of mood scores—how they change and evolve over time—remains less well understood.

To address this gap, we developed a novel pipeline, the Linear Analysis Mixed Model Phenotyper (LAMP), which smooths individual trajectories via a linear mixed model and categorizes them into subgroups, capturing population heterogeneity better than conventional growth models. These subgroups informed trajectory-based phenotypes, such as “Stable/Recovering” and “Resistant/Worsening.” Multivariate logistic regression then explored associations between clinical factors and these trajectory phenotypes. Our framework offers a more precise understanding of the long-term effects of medications (e.g., polypharmacy, psychiatric treatments, and their combinations) as well as other factors (such as substance use disorders, comorbidities, and demographics) on mood symptoms in bipolar disorder. This study also provides valuable insights that could inform and enhance personalized clinical management strategies for BD.

## 2. Methods

### 2.1 Data collection

#### 2.1.1 Participants

We utilized longitudinal data from the Heinz C. Prechter Bipolar Research Project (PrBP), initiated in 2005 at the University of Michigan (McInnis et al., 2018; Yocum et al., 2023). This ongoing study features periodic participant enrollment and dropout for various reasons. PrBP’s extensive phenotypes, assessed through self-reports and clinical evaluations, make it a valuable resource for studying BD and its treatments. Data collected up to December 2, 2022, were included.

The cohort comprises 1,111 individuals: 731 with BD, 23 with Major Depressive Disorder, 34 with other mood disorders, 46 with non-mood psychiatric illnesses, and 277 healthy controls. Our study focused exclusively on BD patients diagnosed according to DSM-IV criteria, distinguishing BP1 (manic episodes) and BP2 (recurrent depression and hypomania). Healthy controls were screened to exclude psychiatric history.

#### 2.1.2 Inclusion and Exclusion Criteria

The study initially included 799 BD patients (BP1 and BP2) with available mood and medication data. Specifically, 747 patients had complete PHQ-9 data, 682 patients had GAD-7 data, and 743 patients had ASRM data. To ensure the analysis focused on the effects of medications during recorded periods, mood data were considered only if they were available between the first recorded medication use date and up to one year after the last recorded medication date. Further refinement required a minimum of three observation points for mood data. Finally, medication data were included only if they were recorded within one year prior to the first mood observation date and up to the last mood observation date, yielding a final sample of 675 patients for PHQ-9, 594 patients for GAD-7, and 670 patients for ASRM (Figure S1).

#### 2.1.3 Outcome

Our primary outcome variables consisted of three self-reported scores measured longitudinally: PHQ-9 (Kroenke et al., 2001), GAD-7 (Spitzer et al., 2006), and ASRM (Altman et al., 1997). The PHQ-9, aligned with DSM-IV criteria, assesses nine depressive symptoms. Each symptom is scored from 0 (“Not at all”) to 3 (“Nearly every day”), yielding a total score range of 0 to 27. The severity of depression is categorized as none (0-4), mild (5-9), moderate (10-14), moderately severe (15-19), and severe (20-27). The GAD-7, also grounded in DSM-IV criteria, evaluates seven anxiety-related symptoms. Items are rated on a scale from 0 to 3, with total scores ranging from 0 to 21. The cut-off scores for categorizing anxiety severity are 5 (mild), 10 (moderate), and 15 (severe). The ASRM, designed to quantify manic symptoms, consists of five items addressing various aspects of mania. Each item is rated on a scale from 0 (no symptom) to 4 (severe symptom), leading to a total score between 0 and 20. Scores above 5.5 on the ASRM are indicative of manic episodes (Miller et al., 2009).

#### 2.1.4 Exposures and Covariates

Exposures were grouped into demographics, medication use, substance use disorders, and comorbidities. In the PrBP, medications are cataloged using the RxNorm Concept Unique Identifier RXCUI (Nelson et al., 2011), a system predominantly utilized in the United States. We have methodically mapped RxNorm to the Anatomical Therapeutic Chemical (ATC) Classification System (World Health Organization, n.d.), employing the OMOP CONCEPT_ANCESTOR table (Observational Health Data Sciences and Informatics, n.d.), with detailed methodology described in the Supplementary Material Section S1.

The ATC system enjoys global application and organizes drugs into a five-level hierarchical structure. Our analysis mainly focuses on the third level — the pharmacological subgroup. Of the 3,797 original RXCUIs in the PrBP database, 843 have been successfully mapped to ATC level three codes pertinent to the nervous system (ATC code “N” and its subcodes). This includes the five most prevalent medication categories among PrBP participants: mood stabilizers, consisting of antiepileptics and lithium (N03A + N05AN01); antipsychotics (N05A, excluding N05AN01); antidepressants (N06A); and anxiolytics (N05B). We calculated both the average number of concurrent medications and the usage frequency for each of these classes across observed time points for each participant.

In accordance with DSM-5 criteria, we have merged “abuse” and “dependence” into a single category termed “substance use disorder” for each substance. This modification has been applied to substances such as alcohol, cannabis, and cocaine, each represented by a binary covariate with “No” as the reference group. For comorbidities, our analysis incorporates binary variables for diagnoses such as PTSD, head injury, migraine, and overweight status, all with “No” as the reference. Additionally, we examined demographic covariates, including gender, age, and marital status at study entry. Gender is categorized into male and female, with “Female” designated as the reference group. Marital status is divided into three categories: “Married”, “Never Married”, and “Divorced/Separated/Widowed”, with the latter category used as the reference group.

### 2.2 Statistical analysis

#### 2.2.1 Descriptive Statistics

After data preprocessing, we summarized demographic characteristics, medication use, substance use disorders, and comorbidity profiles across BP1, BP2, and healthy control cohorts. Healthy controls were used for baseline comparisons only, and is not the focal point of our primary analyses. The differences among the three cohorts were assessed using chi-square tests for categorical variables and ANOVA for continuous variables.

#### 2.2.2 Analysis Pipeline

The primary analysis pipeline, illustrated in Figure 1, consists of four steps, each applied to different mood score types (detailed modeling and term definitions are provided in Supplementary Material Sections S2 and S3).

- <u>Step1</u>: In the first step, we smooth noisy long-term longitudinal mood trajectories into linear trajectories to enhance the interpretability of the analysis. This is achieved using a linear mixed model (Raudenbush and Bryk, 2010) as follows: 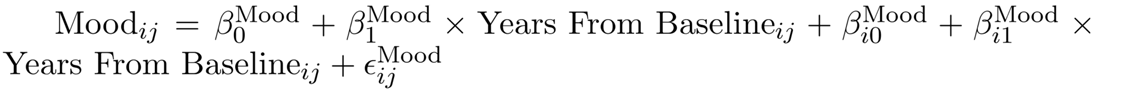 Where 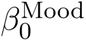 represents population average baseline mood score, and 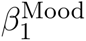 indicates the population average trend of the mood score over time. The terms 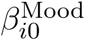 and 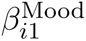 are individual random effects that account for baseline and trend deviations, respectively. For an individual *i*, the estimated baseline of the trajectory is given by 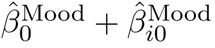, and the estimated trend or slope is 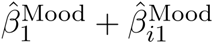, both determined using Best Linear Unbiased Prediction (BLUP; Henderson, 1975).
- <u>Step 2</u>: In the second step, the smoothed trajectories are categorized into subgroups along two dimensions: baseline severity (e.g., within the PHQ-9 framework: low, mild, moderate, or severe depression) and the trajectory’s linear trend, determined by the estimated slopes (recovering: negative slope, static: near-zero slope, or worsening: positive slope). These assessments are conducted separately for each bipolar patient using the PHQ-9, GAD-7, and ASRM scores.
- <u>Step 3</u>: In the third step, we define a binary phenotype “Stable/Recovering” and “Resistant/Worsening” for both individual and aggregated scores. For individual scores (PHQ, GAD, or ASRM), “Stable/Recovering” refers to individuals with either static scores and a low baseline severity or those with recovering scores. Conversely, “Resistant/Worsening” refers to individuals with either static scores and a moderate or high baseline severity or those with worsening scores. The integrated score follows similar criteria.
- <u>Step 4</u>: In the final step, multivariate logistic regression analyses were conducted to examine the associations between covariates of interest and the binary phenotype (as defined in Step 3) across the PHQ-9, GAD-7, ASRM, and the integrated score, separately within the BP1 and BP2 cohorts. Additionally, interactions between mood stabilizers, antipsychotics, and antidepressants were evaluated in separate models, with the significance of these interaction terms tested using likelihood ratio tests (LRTs).

**Figure 1:**
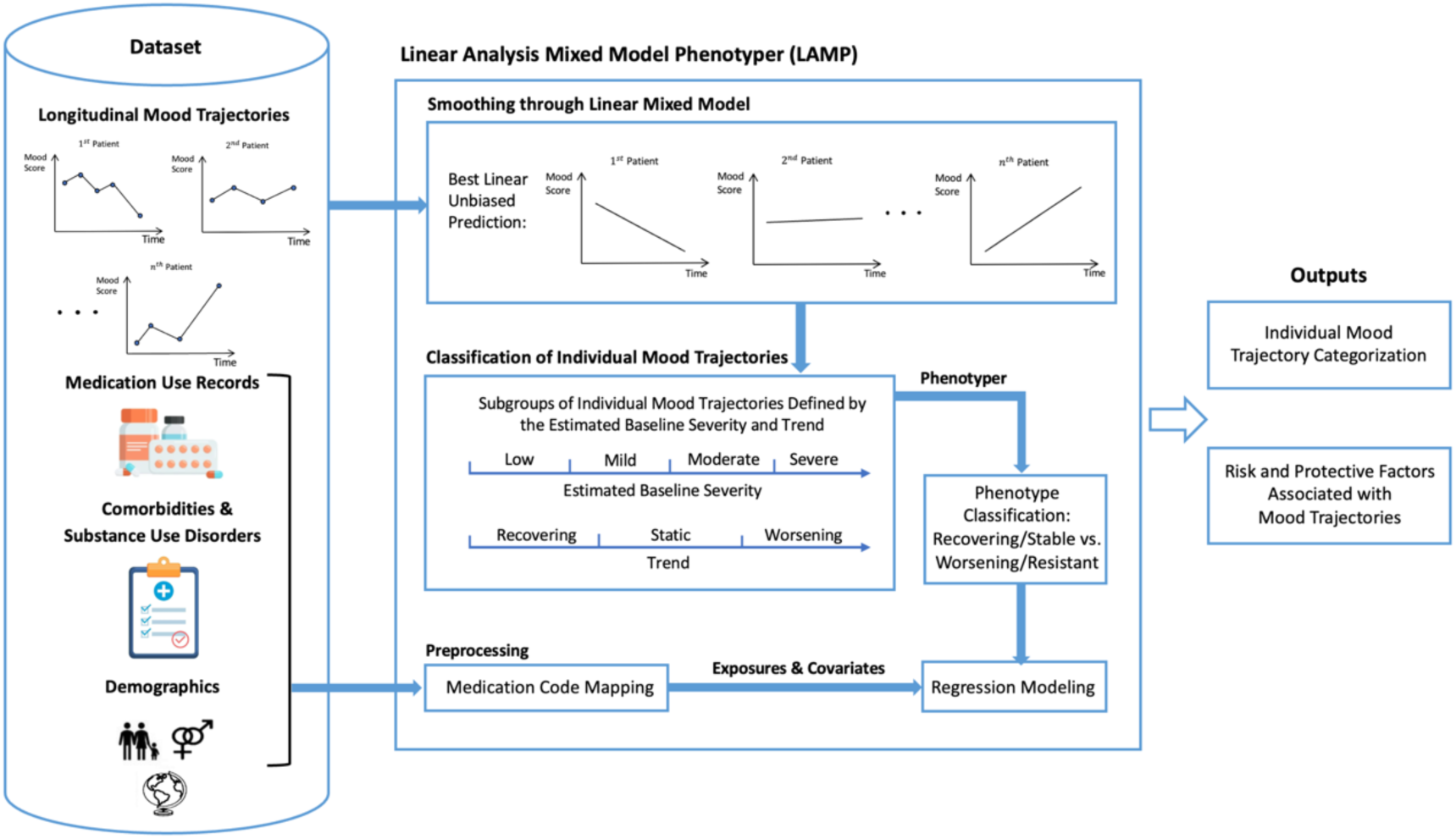
Overview of the LAMP modeling pipeline for examining different patterns in mood trajectories and the impact of exposures on them. The pipeline includes smoothing longitudinal mood trajectories using a linear mixed model, classifying trajectories into phenotypes based on baseline severity and trends, and analyzing the influence of exposures (medications, comorbidities, and substance use) on these phenotypes through logistic regression models. Outputs include mood trajectory classifications and statistical inferences about exposure effects.

## 3. Results

### 3.1. Descriptive statistics

As shown in Table 1, the final analytical sample comprised 525 participants from the BP1 cohort and 150 from the BP2 cohort. All included participants had data for the PHQ outcomes. However, 70 BP1 and 11 BP2 patients lacked GAD outcomes, primarily because the GAD data collection commenced in 2012, whereas the study began in 2005. Additionally, 5 BP1 patients did not have ASRM outcomes. The summary statistics of healthy control group are provided in Table S1.

**Table 1:**
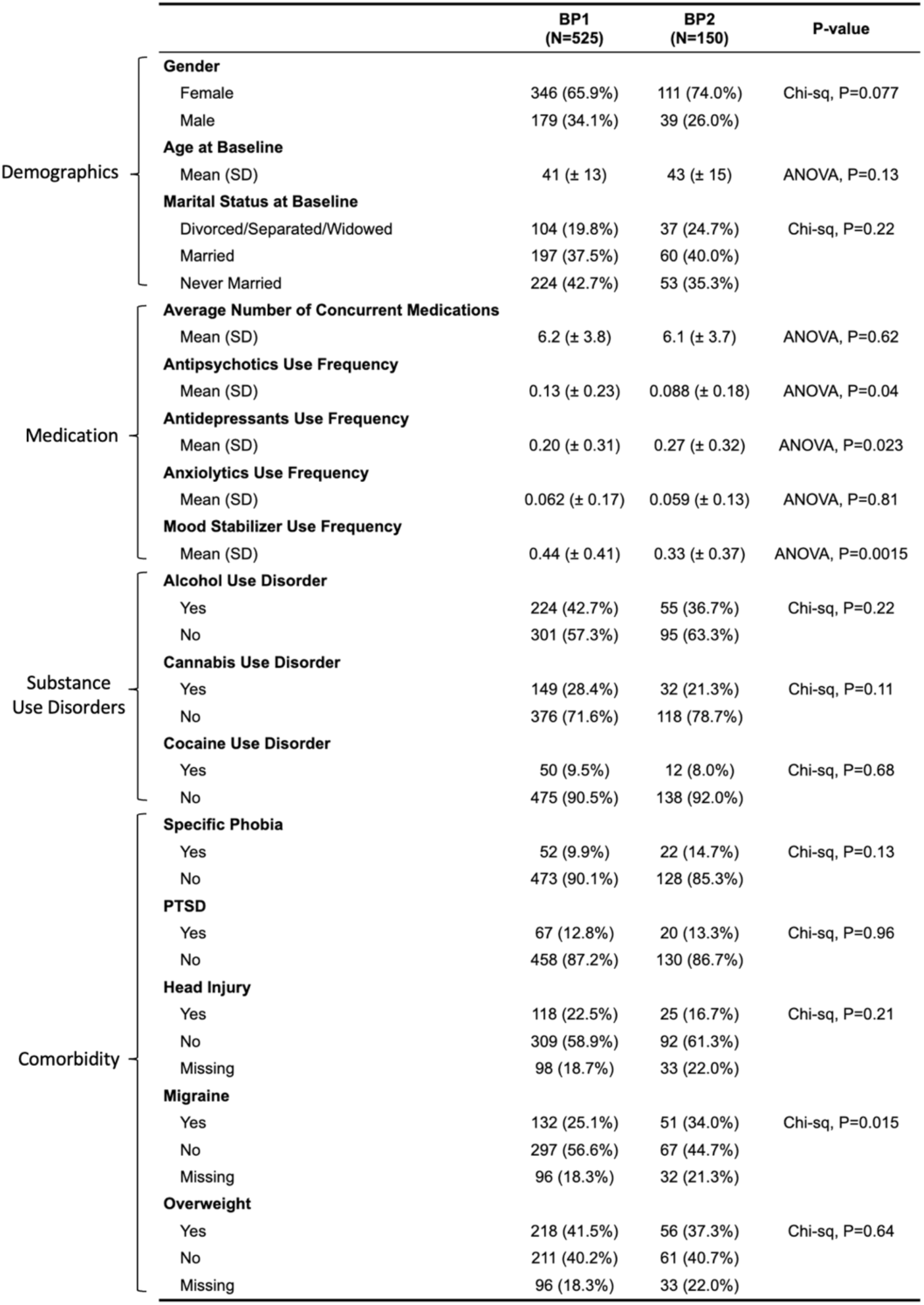
Summary statistics for the BP1 and BP2 cohorts. For continuous covariates, means and standard deviations are provided. For categorical variables, sample sizes and corresponding proportions are provided. P-values compare BP1 and BP2 using two-sided t-tests (continuous) or chi-square tests (categorical).

The majority of the sample was female, with females comprising 66% of the BP1 cohort and 74% of the BP2 cohort. The average age at study entry was approximately 40 for each cohort. The average number of concurrent medications for individuals in the BP1 and BP2 cohorts was approximately 6. This indicates that, on average, individuals in the BP1 and BP2 cohorts are experiencing polypharmacy. Additionally, the use frequencies of antipsychotics, antidepressants, mood stabilizers, and the presence of migraines, as outlined in Table 1, differed significantly between the BP1 and BP2 cohorts, as determined by chi-square tests and ANOVA.

### 3.2 Classification of Mood Trajectories

#### 3.2.1 Analysis of Individual Mood Scores

The mood trajectories for BP1 and BP2 patients (Figure S2) show distinct patterns. To characterize these trajectories, they were smoothed and stratified into subgroups based on baseline severity levels and trends (Figure 2). The trends, represented by trajectory slopes, indicate the average annual change in mood scores, while baseline severity is captured by intercepts. Table S2 provides sample sizes, average intercepts, and slopes for each subgroup.

**Figure 2:**
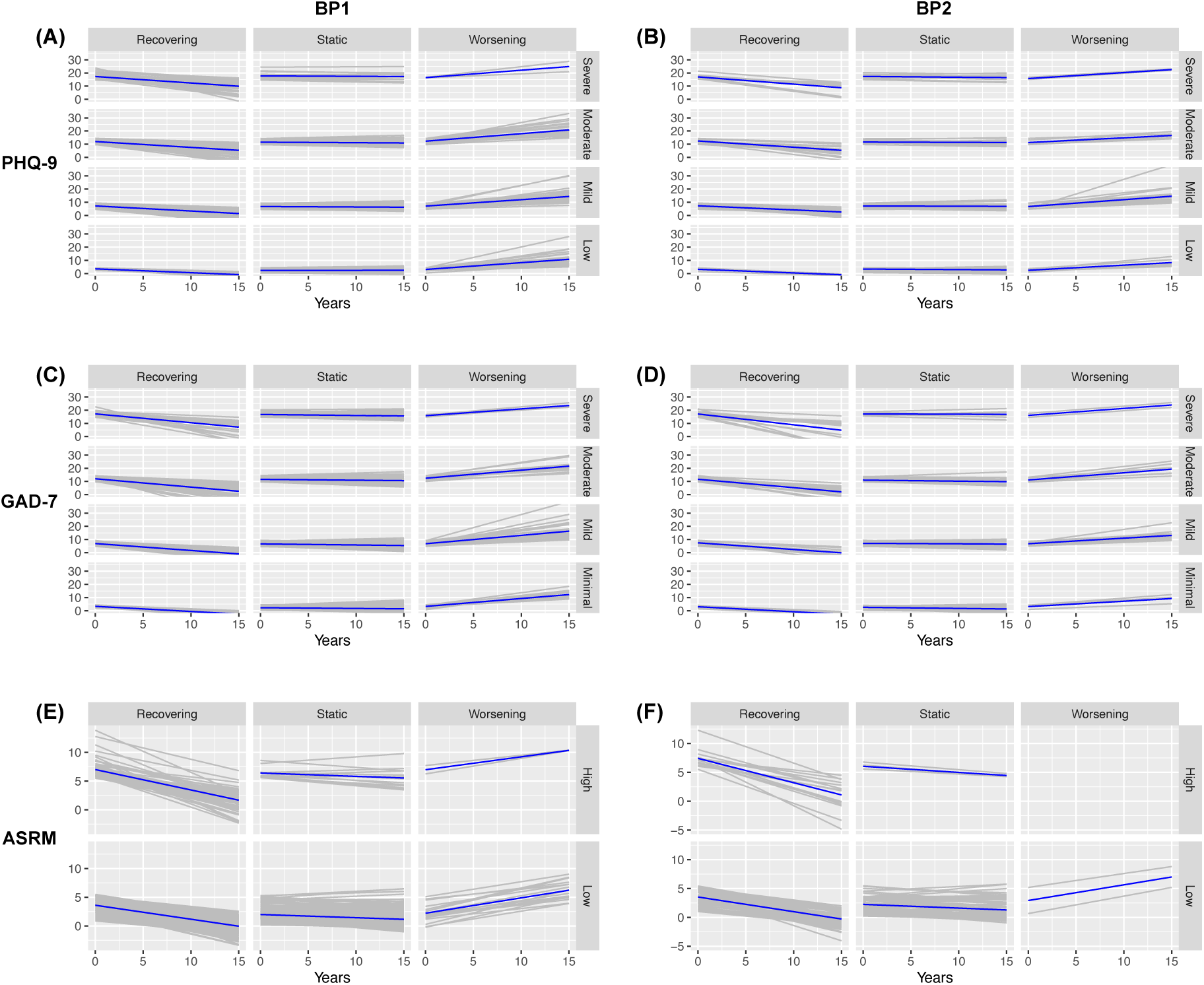
Visualization of Smoothed Mood Trajectories. Panels A, C, and E display PHQ-9, GAD-7, and ASRM trajectories for BP1, respectively. Panels B, D, and F represent corresponding trajectories for BP2. Subgroups are categorized by baseline severity and progression trends.

In Figure 2, static subgroups show minimal slope changes, while recovering and worsening subgroups display clear improvement or deterioration trends. Over a decade, worsening subgroups’ scores increased by approximately 5 points for PHQ-9 and GAD-7 and approximately 3 points for ASRM, while recovering subgroups decreased by similar magnitudes. Fewer participants in the ASRM worsening subgroup suggest a generally more stable ASRM trend compared to PHQ-9 and GAD-7. Figure S3 shows density distributions of intercepts and slopes by mood score type, revealing similar trends between BP1 and BP2, though BP2 patients tend to have slightly higher baseline scores. Figure S4 shows original and smoothed trajectories for the healthy control group, most of whom fall into low to mild baseline severity categories.

#### 3.2.2 Integrated Analysis of Mood Scores

Based on the classification of mood trajectories outlined in the previous section, we explored the intersections of symptom trend categories—Recovering, Static, and Worsening—across the three mood score assessments within the BP1 and BP2 cohorts. Figure 3 shows that 90 (22%) of BP1 and 18 (16%) of BP2 patients exhibit static trends across PHQ-9, GAD-7, and ASRM scores, representing the largest subgroup in each cohort. Additionally, 52 (13%) of BP1 and 15 (13%) of BP2 patients demonstrate a recovering trend across these mood scores, making this the third largest group. Notably, the number of individuals experiencing at least one worsening mood score is generally smaller than those who remain static or are recovering across all mood scores, suggesting partial effectiveness of treatments for bipolar disorder. In the next section, we combined these three types of mood trajectories to define an integrated “Stable/Recovering” versus “Resistant/Worsening” mood trajectory. Unlike PHQ-9, ASRM, or GAD-7, which assess individual dimensions of mood (depression, mania, or anxiety), this integrated measure reflects a broader aspect of mood status by synthesizing patterns across all three domains into a single classification.

**Figure 3:**
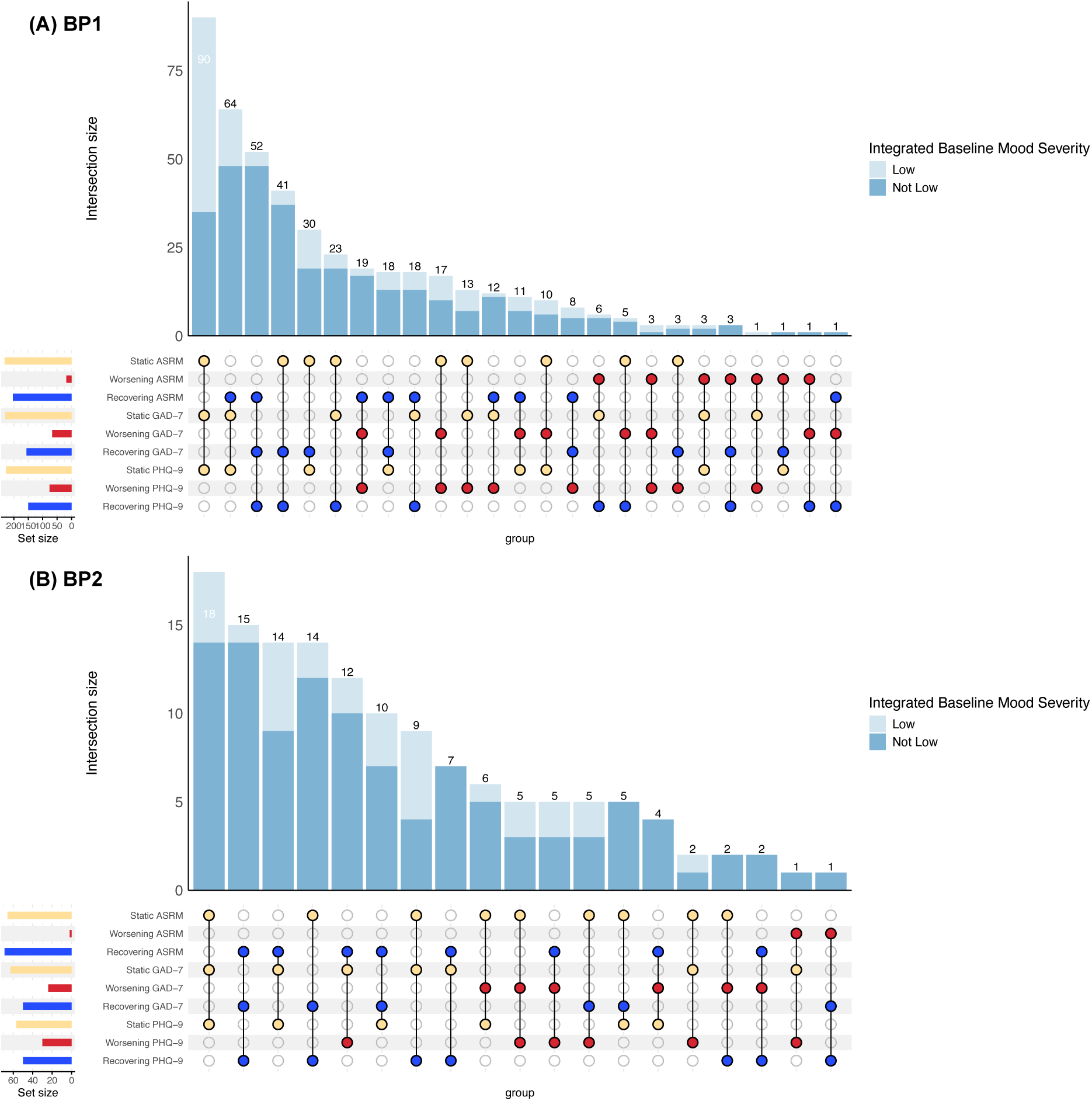
Upset plot analysis with stacked barplots for BP1 (Panel A) and BP2 (Panel B) patients with complete PHQ-9, GAD-7, and ASRM data. This visualization explores intersections of mood state categories—Recovering, Static, and Worsening—across the three assessments. The light blue and blue colors in the stacked bars correspond to the “low” and “not low” integrated baseline mood severity. The ‘Low’ group is defined such that individuals must have all baseline severity scores categorized as low for PHQ-9, GAD-7, and ASRM, or at most one score classified as mild. All other individuals are classified as ‘Not Low’.

### 3.3 Relationship Between Mood Trajectories and Contributing Factors

#### 3.3.1 Descriptive Analysis of the Two Phenotype Groups

The phenotype of mood trajectories, as defined in Section 2.2, is binary, consisting of the categories “Stable/Recovering,” considered as a positive trajectory, and “Resistant/Worsening,” viewed as a negative trajectory. As shown in Table S3 and S4, the distribution for “Stable/Recovering” vs. “Resistant/Worsening” includes 277 vs. 248 BP1 patients and 67 vs. 83 BP2 patients for PHQ-9; 256 vs. 199 BP1 patients and 67 vs. 72 BP2 patients for GAD-7; 489 vs. 31 BP1 patients and 144 vs. 6 BP2 patients for ASRM; and 238 vs. 177 BP1 patients and 65 vs. 49 BP2 patients for the integrated mood measure. Due to the limited number of BP2 patients (only 6) in the “Resistant/Worsening” group for ASRM, statistical inference analysis was not conducted for ASRM in BP2.

#### 3.3.2 Statistical Examination of the Relationships

Multivariate logistic regression analyses were performed to examine the associations between covariates of interest and the binary phenotype, where “Stable/Recovering” was coded as reference. The regression results are presented in Figure 4 as forest plots, showing odds ratios along with confidence intervals and P-values. Panel A displays the results for BP1, and panel B for BP2. Significant associations (P-value < 0.05) are highlighted in red. Baseline mood score, referred to as “Baseline,” was used as an adjustment for each mood score analysis. For the integrated analysis, the integrated baseline mood severity score was used for adjustment.

**Figure 4:**
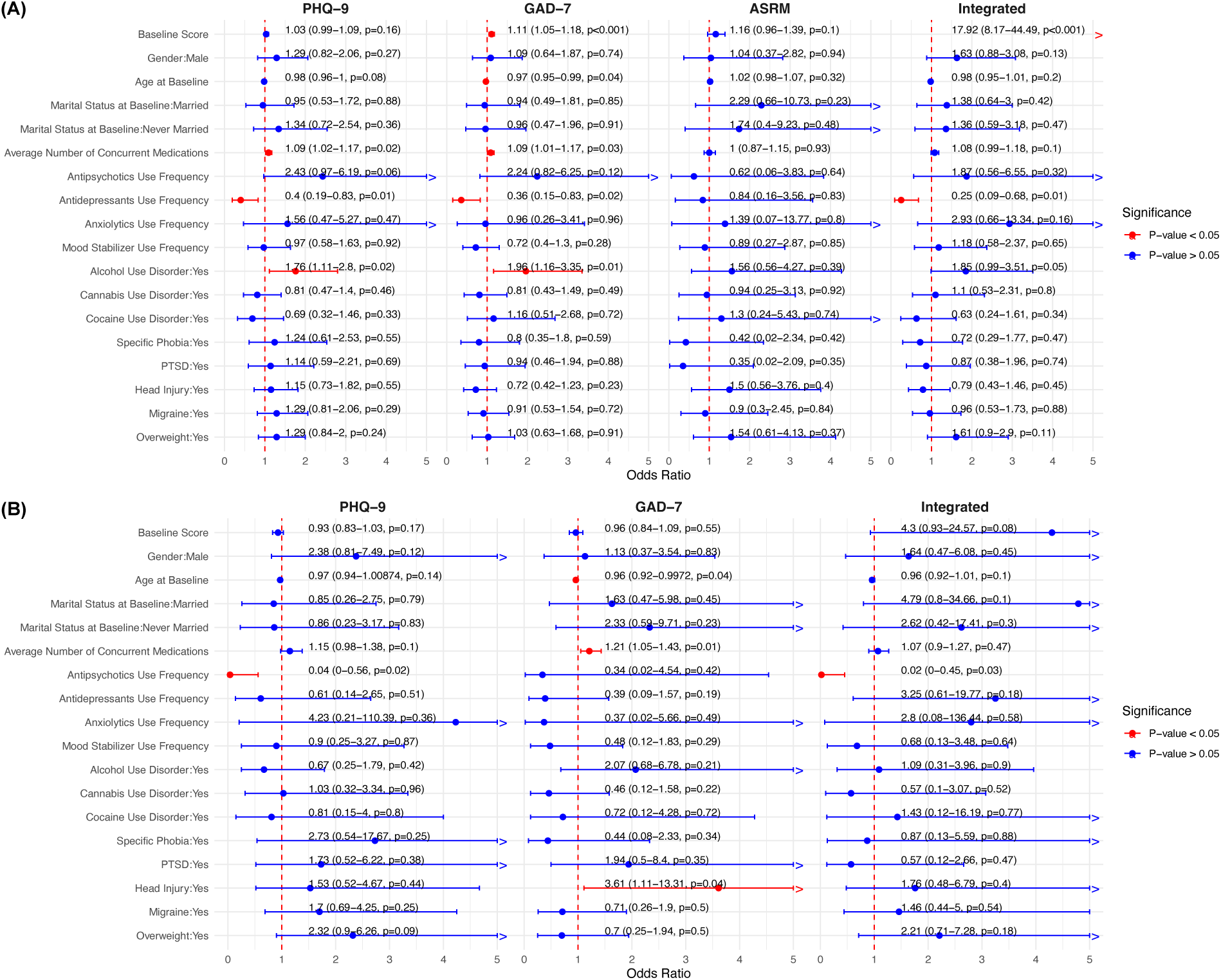
Forest plots showing odds ratios (ORs) and 95% confidence intervals for covariate associations with “Stable/Recovering” and “Resistant/Worsening” trajectory groups for BP1 (Panel A) and BP2 (Panel B). Each column corresponds to PHQ-9, GAD-7, ASRM, and their integrated phenotype. Covariate names and reference groups are listed on the left. Points represent OR estimates, and bars show confidence intervals. The right side of bars that extend beyond a value of 5 are marked with a ‘>’ for clearer visualization. Text annotations above each bar provide detailed statistics: odds ratio, confidence interval, and p-value. An OR less than 1 indicates a tendency towards the “Stable/Recovering” group, while an OR greater than 1 indicates a tendency towards the “Resistant/Worsening” group. Significant associations (P-value < 0.05) are highlighted in red within the forest plots.

##### Concurrent Medication Use

The number of concurrent medications was significantly associated with more resistant/worsening trajectories in both BP1 and BP2. In BP1, each additional medication was associated with 9% higher odds of being in the worse trajectory group on the PHQ-9 (Odds Ratio: 1.09, CI: 1.02–1.17, p = 0.01) and the GAD-7 (Odds Ratio: 1.09, CI: 1.01–1.17, p = 0.03). Similarly, in BP2, each additional medication was associated with 21% higher odds of being in the worse trajectory group on the GAD-7 (Odds Ratio: 1.21, CI: 1.05–1.43, p = 0.01), indicating that patients requiring multiple medications may have more severe or complex illness.

##### Antipsychotic Use

Antipsychotic use in BP2 was significantly associated with a stable/recovering trajectory on the PHQ-9 (Odds Ratio: 0.04, CI: 0.00–0.56, p = 0.02) and the integrated mood measure (Odds Ratio: 0.02, CI: 0.00–0.45, p = 0.03), suggesting their potential effectiveness in managing symptoms in BP2 patients. These findings highlight the potential benefits of antipsychotics in improving mood trajectories for BP2. In contrast, no significant associations were observed in BP1.

##### Antidepressant Use

Antidepressants were consistently associated with more favorable disease trajectories in BP1. Antidepressant use was significantly linked to a more stable/recovering trajectory across the PHQ-9 (Odds Ratio: 0.40, CI: 0.19-0.83, p=0.01), GAD-7 (Odds Ratio: 0.36, CI: 0.15-0.86, p=0.02), and the Integrated mood measure (Odds Ratio: 0.25, CI: 0.09-0.68, p=0.01). No significant associations were observed for antidepressant use in BP2, suggesting that its effects may be more pronounced in BP1 patients.

##### Substance Use Disorders

Alcohol use disorder was consistently associated with resistant/worsening trajectories in BP1. Significant associations were observed with worse trajectories on the PHQ-9 (Odds Ratio: 1.83, CI: 1.15-2.93, p=0.01), GAD-7 (Odds Ratio: 1.96, CI: 1.16-3.35, p=0.01), and the Integrated mood measure (Odds Ratio: 1.85, CI: 0.99-3.51, p=0.05). However, no significant associations between alcohol use disorder and mood scores were found in BP2.

##### Comorbidity

In BP2, head injury was significantly associated with a more resistant/worsening trajectory on the GAD-7 (Odds Ratio: 3.61, CI: 1.11-13.31, p=0.04). This suggests that comorbid conditions such as head injury may complicate the management of anxiety symptoms in BP2 patients.

##### Demographics

Older age of onset was significantly associated with a more stable/recovering trajectory on the GAD-7 in BP1 (Odds Ratio = 0.97, CI: 0.95–0.99, p = 0.04) and BP2 (Odds Ratio = 0.96, CI: 0.92–1.00, p = 0.04), suggesting that older patients have a higher response to medication compared to younger patients. Marital status was not significantly associated with mood scores in either group.

#### 3.3.3 Impact of Psychiatric Medication Interactions on Trajectory Outcomes

To examine the long-term impact of interactions between the use of antidepressants, antipsychotics, and mood stabilizers on mood scores, LRTs were conducted for each regression model in Section 3.3.2 to assess the significance of including these interaction terms. The results indicated that the interaction between antidepressants and antipsychotics was significant in BP1 for both the PHQ-9 (LRT: p = 0.00059) and the integrated analysis (LRT: p = 0.00014). For PHQ-9, increasing antidepressant and antipsychotic use from 0% to 20% raised the odds of resistant/worsening trajectories by 2.09 times (95% CI: 1.33– 3.28, p = 0.0013). Similarly, for the integrated mood score, the odds increased by 3.06 times (95% CI: 1.63–5.72, p < 0.001). No significant interaction was found between antidepressant use and mood stabilizers.

To further illustrate these results, we categorized individuals into three groups: (1) frequent antidepressant use only, (2) frequent combined use of antidepressants and antipsychotics, and (3) neither (for detailed definitions, refer to Supplementary Material S3). For each group, we calculated the proportion of individuals in the “Stable/Recovering” and “Resistant/Worsening” categories. Figure 5 shows that individuals with frequent use of both antidepressants and antipsychotics were significantly more likely to be in the resistant/worsening group for PHQ-9 compared to those with frequent antidepressant use only (Pearson’s Chi-square test, p = 0.0063). A similar analysis was conducted for mood stabilizer use versus combined mood stabilizer and antidepressant use, with no significant results detected. Additionally, Figure S5 indicates that individuals with frequent use of all three medication types (antidepressants, antipsychotics, and mood stabilizers) were more likely to be in the resistant/worsening group for PHQ-9 compared to the other two groups, although this trend did not reach statistical significance.

**Figure 5.**
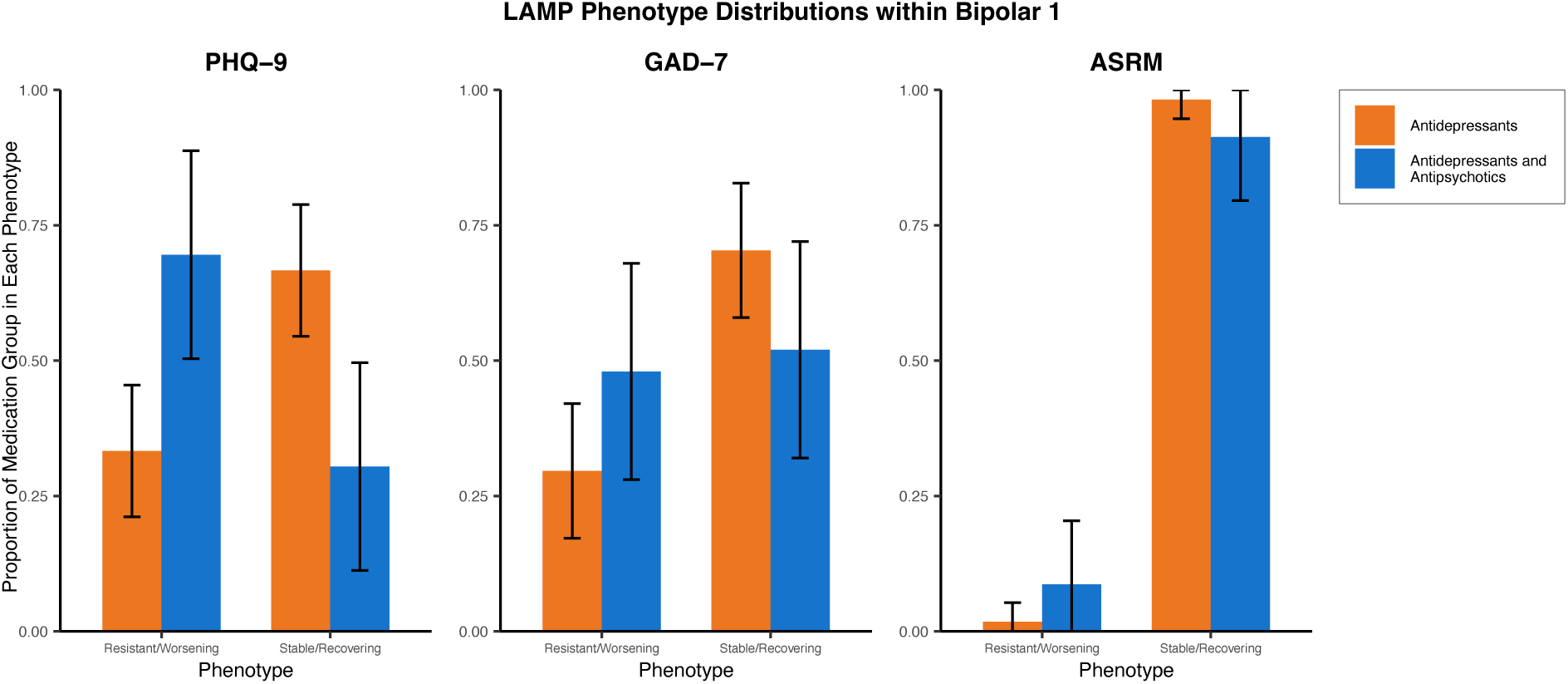
LAMP phenotype distributions in BP1 for PHQ-9, GAD-7, and ASRM. Orange bars represent frequent antidepressant use, while blue bars show combined use of antidepressants and antipsychotics. Error bars indicate 95% confidence intervals.

## 4. Discussion

Biological signals from EHR data or surveys often suffer from noise and inconsistencies due to variable acquisition intervals and baselines (Holmes et al., 2021). Using data from PrBP, LAMP addresses this issue by employing a mixed model to condense noisy and variable signals into a fixed set of interpretable parameters and groups, facilitating analysis while retaining biologically important context. Compared to traditional methods like Latent Class Growth Analysis (LCGA; Jung and Wickrama, 2008), which often oversimplifies mood trajectories into 3–4 classes, LAMP provides a more nuanced framework that accommodates the complexity of mood trajectories, such as the 12 potential classes from PHQ-9 data (see Supplementary Material, Section S4, for a detailed comparison with LCGA).

Antidepressant use in bipolar disorder remains contentious and appears to differ by subtype. In our long-term analysis, antidepressant exposure was associated with more favorable depressive trajectories in BP1 but not in BP2, which is consistent with consensus statements that antidepressants in BP2 are generally well tolerated yet have uncertain efficacy for acute depression (Pacchiarotti et al., 2013). If antidepressants are used in BP1, clinical guidelines advise co-prescription with a mood-stabilizing agent because of potential mood switching, although estimates of switch risk vary across studies (Melhuish Beaupre et al., 2020; Pacchiarotti et al., 2013; Sachs et al., 2007). Our cohort reflects this practice pattern: mood stabilizer use was significantly more frequent in BP1 than BP2 (44% vs. 33%). In addition, we did not observe a significant long-term increase (or decrease) in manic outcomes when antidepressants were used adjunctively with mood stabilizers. These findings support the view that antidepressants in bipolar disorder should neither be universally endorsed nor categorically avoided, but rather considered on a case-by-case basis (Pacchiarotti et al., 2013).

Building on these subtype-specific differences, we also observed that antipsychotic use was associated with improved mood trajectories in BP2, consistent with existing evidence supporting their role in bipolar depression management (Rybakowski, 2023). In contrast, BP1 presents a different pattern: while antidepressants consistently lead to better trajectories, the combination of antidepressants and antipsychotics may have adverse effects over time. This does not contradict evidence of short-term agent-specific benefits— for example, the olanzapine–fluoxetine combination demonstrated modest acute efficacy compared to olanzapine alone (Hu et al., 2022)—but rather highlights that long-term effects at the class level may differ from short-term trial results. Another key finding from our study was the high prevalence of polypharmacy (mean ≈6 concurrent medications in PrBP), which was associated with more resistant or worsening trajectories in both BP1 and BP2. This observation aligns with previous research showing that complex polypharmacy is correlated with higher rates of depression and comorbid anxiety disorders. (Weinstock et al., 2014). Clinically, the use of multiple medications and higher doses often reflects the degree of treatment resistance, characterized by symptoms that fail to respond to conventional treatment. These findings underscore the importance of further research into personalized treatment strategies, considering differences between bipolar subtypes, medication classes, treatment approaches and short-term versus long-term mood trajectories, as well as the risks of polypharmacy.

Our finding that alcohol use disorder was associated with worsening mood trajectories in BP1 aligns with previous studies linking worsening mood trajectories to alcohol use disorders (Smith et al., 2025; Sperry et al., 2024). Similarly, the significant association between head injury and worsening GAD-7 trajectories in BP2 reflects the well-documented connection between head injury and bipolar disorder; individuals with traumatic brain injury are estimated to be 28 times more likely to develop bipolar disorder (Altuwairqi, 2023). These findings underscore the critical impact of substance use disorders and comorbid conditions on mood symptom management in bipolar patients.

Although many results did not show significance, we cannot conclude that they lack effect. The non-significance may be due to the relatively small sample size, especially for BP2, the complexity of mood trajectories, the retrospective nature of the data, and potential confounders not accounted for in our analysis. Our focus on long-term effects, despite psychiatric medications typically having short-term impacts, may also reflect different findings with short-term effects, given the chronic nature of bipolar disorder. Future research should aim for larger sample sizes and more precise mood measurements to better capture long-term effects. Investigating moderators and mediators of mood trajectories and employing longitudinal studies with frequent assessments could uncover additional trends and enhance our understanding and management of bipolar disorder.

While LAMP provides a valuable framework for analyzing longitudinal trajectories, it has inherent limitations. One significant challenge is the uncertainty surrounding the sequential effects of medications due to ambiguity in their prescribed order—antidepressants may have been initiated first, followed by antipsychotics, or vice versa, or both may have been prescribed simultaneously. This ambiguity complicates the assessment of causal relationships. However, with 17 years of data in our study, the consistent worsening trajectories observed in individuals on both treatments suggest a potential link between certain medication combinations and long-term effects. On the other hand, the nature of the data makes it difficult to accurately assess short-term effects. Additionally, the linearity of Linear Mixed Models presents another limitation. The complexity of mood and medication data in the PrBP cohort can lead to inaccuracies if non-linear relationships, such as quadratic trends, are not considered. Long-term linear trends may also fail to capture the dynamic fluctuations of mood over time. Lastly, like many methodologies, the effectiveness of LAMP depends heavily on the quality of the data it is applied to. In the case of the PrBP cohort, limitations such as geographic and racial imbalances among participants (Yocum et al., 2023) must be acknowledged.

## 5. Conclusions

This study demonstrates the effectiveness of LAMP in identifying distinct mood trajectory subgroups in bipolar disorder using 17 years of data from the Prechter Bipolar Research Program. Key findings include associations of polypharmacy and alcohol use disorder with increased risk of resistant or worsening trajectories, beneficial effects of antidepressants in BP1 negatively impacted by concurrent antipsychotic use, and improved trajectories associated with antipsychotic treatment in BP2. These results underscore the importance of individualized medication strategies, careful management of polypharmacy and medication combinations, and targeted interventions addressing substance use disorders and comorbid conditions. Future research should focus on refining data integration, medication tracking, and mood assessments to further enhance clinical outcomes.

## Data Availability

The data that support the findings of this study are available from the Heinz C. Prechter Bipolar Research Program but restrictions apply to the availability of these data, which were used under license for the current study, and so are not publicly available. Data are however available from the authors upon reasonable request and with permission of the Heinz C. Prechter Bipolar Research Program. The code to replicate all results presented in this paper, along with the RXCUI-to-ATC mapping table and tutorial example code, is available at https://github.com/qingzliu/LAMP.

## Declarations

## Ethics Approval and Consent to Participate

The study was conducted in accordance with the Declaration of Helsinki, and the protocol was approved by the Ethics Committee of the University of Michigan (IRB approval numbers: HUM00000606 and HUM00231342). All participants provided written informed consent.

## Conflict of Interests

Brian Athey is the Co-Founder and Chief Science and Technology Officer and Alex Ade is the Systems Research Programmer at Phenomics Health Incorporated (Plymouth Township, MI). They both have a Conflict-of-Interest Management Plan on file with the University of Michigan, approved by the Regents of the University of Michigan. These affiliations may present a potential conflict of interest related to this work. All relevant financial and professional relationships have been disclosed, and appropriate measures are being taken to manage or mitigate any potential conflicts in accordance with institutional policies.

## Funding

This work was supported by Phenomics Health Incorporated (Plymouth Township, MI; Contract NO36672). Additional support was provided by Training Grant T32 GM141746 from the National Institutes of Health, the Department of Computational Medicine and Bioinformatics at the University of Michigan Medical School, and the Heinz C. Prechter Bipolar Research Fund. The content is solely the responsibility of the authors and does not necessarily represent the official views of the funding sources.

## Authors’ Contributions

Qingzhi Liu: Conceptualization, Methodology, Data curation, Software, Formal analysis, Validation, Writing - Original Draft, Visualization

Pranjal Srivastava: Methodology, Validation, Formal analysis, Software, Writing - Original Draft, Writing - Review & Editing

Ivan Rozhkov: Methodology, Validation, Software, Formal analysis, Writing - Original Draft, Writing - Review & Editing

Gregory A. Farnum: Conceptualization, Methodology, Supervision, Writing - Review & Editing

Monica J. Holmes: Conceptualization, Writing - Review & Editing

Lars Fritsche: Conceptualization, Methodology

David Belmonte: Conceptualization, Methodology, Writing - Review & Editing

Alex Ade: Data resources, Data curation, Software

Anastasia Yocum: Data resources, Data curation

Melvin Mclnnis: Conceptualization, Review & Editing, Funding acquisition

Veera Baladandayuthapani: Conceptualization, Methodology, Formal analysis, Supervision, Writing - Review & Editing, Project Administration

Brian D. Athey: Conceptualization, Methodology, Supervision, Writing - Review & Editing, Project administration, Funding acquisition

## Acknowledgments

We would like to thank all the participants of the Heinz C. Prechter Bipolar Research Program for their invaluable contributions to this study.

## Declaration of Generative AI and AI-Assisted Technologies

The authors used OpenAI’s ChatGPT to assist in the preparation of this manuscript. Specific prompts and guidance were provided to the tool, and the authors subsequently reviewed and edited all output to ensure accuracy, clarity, and integrity.

## Notes

### Author Declarations

Ethics Committee/IRB of the University of Michigan gave ethical approval for this work (IRB approval numbers: HUM00000606 and HUM00231342)

