## Supplementary Materials for "A Linear Analysis Mixed Model Phenotyper (LAMP) for Bipolar Disorder: Evaluating the Impact of Medication, Substance Use Disorders, and Comorbidities on Mood Trajectories"

### S1. RXCUI to ATC mapping

In our study, the process of mapping RxNorm Concept Unique Identifiers (RxCUIs) to the Anatomical Therapeutic Chemical (ATC) classification system presented a complex challenge, particularly due to the data being based on ingredients rather than specific drug names. To effectively address this issue, we adopted a modified approach that centers on a more targeted scope and stringent mapping criteria.

We narrowed our focus to ATC Level 5 codes specifically related to the "NERVOUS SYSTEM" category, denoted by codes starting with "N". This decision to concentrate on a single category significantly simplifies the task by avoiding the ambiguities and complexities associated with broader drug categories. In the mapping process, we accepted the existing RxNorm to ATC mappings, with the condition that the original RxCUI was not an ingredient. Additionally, for ingredient-based RxNorm entries, we implemented a strict criterion: an ATC Level 5 code mapping was only accepted if the ingredient in RxNorm was explicitly mentioned in the corresponding ATC name. This approach ensures a more precise and relevant correlation between the RxNorm data and the ATC classification. In the final stage of our analysis, we rolled up the mapped ATC Level 5 codes to all their parent levels.

The rationale behind this methodology is twofold. Firstly, it streamlines the process by focusing on drugs impacting the nervous system, thus reducing potential complexities. Secondly, it allows for context-dependent mapping. Since our participants often refer to drugs by their names or bring their medication packages for reference, we have a reliable context for directly mapping RxNorm codes to ATC codes, as supported by the OMOP concept ancestor table. Additionally, when participants mention only an ingredient, such as "I take lithium," it's more likely they're referring to a medication where that ingredient plays a central role. Our approach, while not flawless, strikes a balance between accuracy and simplicity, and is particularly effective for our study's focus on medications related to the nervous system.

In our study, we focused on specific ATC Level 3 codes, encompassing antiepileptics (N03A), antipsychotics (N05A), antidepressants (N06A), and anxiolytics (N05B). A noteworthy aspect of our analysis is the classification of lithium, which is assigned the

ATC Level 5 code N05AN01. Although lithium is classified under antipsychotics in the ATC system, we treated it as a mood stabilizer for the purposes of our study. Consequently, we excluded N05AN01 from the broader N05A (antipsychotics) category and analyzed it together with antiepileptics as part of the mood stabilizer group (N03A + N05AN01).

### S2. Smoothing and Classification of Mood Trajectories

Our statistical analysis methodology is a structured, multi-step process specifically designed to assess mood trajectories within the Bipolar I (BP1), Bipolar II (BP2), and healthy control cohorts. Focusing on various mood scores including the Patient Health Questionnaire (PHQ), Generalized Anxiety Disorder 7-item (GAD), and Altman Self-Rating Mania Scale (ASRM), this approach provides a comprehensive examination of these trajectories.

In the first phase of our analysis, we smooth individual mood trajectories into linear trends. This is carried out separately for the BP1, BP2, and healthy control cohorts, across different mood score types. Each individual's linear trend is defined by two key parameters: an estimated intercept and slope. For example, in analyzing BP1 patients using the PHQ score, we apply a linear mixed model as follows:

$$\mathrm{PHQ}_{ij} = \beta_0^{\mathrm{PHQ}} + \beta_1^{\mathrm{PHQ}} \times \mathrm{Years\ From\ Baseline}_{ij} + \beta_{i0}^{\mathrm{PHQ}} + \beta_{i1}^{\mathrm{PHQ}} \times \mathrm{Years\ From\ Baseline}_{ij} + \epsilon_{ij}^{\mathrm{PHQ}}$$

The vector $\beta_i^{\mathrm{PHQ}} = [\beta_{i0}^{\mathrm{PHQ}}, \beta_{i1}^{\mathrm{PHQ}}]$ is assumed to follow a normal distribution $N(0_2, \Omega)$, with the PHQ score for individual $i$, $\mathrm{PHQ}_i$, adhering to a normal distribution $N(0_{n_i}, \Sigma)$. This model is similarly employed for GAD and ASRM scores. The estimated intercept of the PHQ trajectory for an individual $i$ is $\hat{\beta}_0^{\mathrm{PHQ}} + \hat{\beta}_{i0}^{\mathrm{PHQ}}$, denoted as $Intercept_i^{\mathrm{PHQ}}$, determined through Best Linear Unbiased Prediction (BLUP). The corresponding estimated slope is $\hat{\beta}_1^{\mathrm{PHQ}} + \hat{\beta}_{i1}^{\mathrm{PHQ}}$, represented as $Slope_i^{\mathrm{PHQ}}$.

In the second phase, we categorize individual mood trajectories based on their post-smoothing linear trends, independently for the BP1, BP2, and healthy control cohorts, and across each mood score type. The baseline severity for each mood score type is classified based on estimated individual intercepts from linear trends. For PHQ, this ranges from 0 to 4.5 for low depression, 4.5 to 9.5 for mild depression, 9.5 to 14.5 for moderate depression, and 14.5 to 27 for severe depression. For GAD, $Intercept_i^{\mathrm{GAD}}$ spans from 0 to 4.5 for minimal anxiety disorder, 4.5 to 9.5 for mild anxiety disorder, 9.5 to 14.5 for moderate anxiety disorder, and 14.5 to 27 for severe anxiety disorder. For ASRM, $Intercept_i^{\mathrm{ASRM}}$ below 5.5 signifies low and above 5.5 as high mania. Linear progression is categorized into recovering (slopes are negative), stable (slopes approximately 0), and worsening (slopes are positive). We define a threshold $\delta$ such that slopes from $-\infty$ to $-\delta$ correspond to recovering, slopes from $-\delta$ to $\delta$ as stable, and slopes from $\delta$ to $\infty$ as worsening. For each mood score type, $\delta$ is determined to ensure that the recovering group comprises 50% of the combined BP1 and BP2 cohorts. In our analysis, the value of $\delta$ was determined to be 0.17 for PHQ-9, 0.28 for GAD-7, and 0.15 for ASRM.

This methodical and systematic approach provides an intricate framework for comprehending the diverse symptom trajectories, thus enhancing both the interpretability and applicability of our findings.

### S3. Definitions of Key Terms

| Term | Definition |
|---|---|
| PHQ Categories | Depression severity based on PHQ-9 scores: 0-4.5 (low), 4.5-9.5 (mild), 9.5-14.5 (moderate), 14.5-27 (severe). |
| GAD Categories | Anxiety severity based on GAD-7 scores: 0-4.5 (minimal), 4.5-9.5 (mild), 9.5-14.5 (moderate), 14.5-21 (severe). |
| ASRM Categories | Mania severity based on ASRM scores: below 5.5 (low), above 5.5 (high). |
| Stable/Recovering (PHQ, GAD or ASRM) | Individuals with either static scores and a low baseline severity or those with recovering scores. |
| Resistant/Worsening (PHQ, GAD or ASRM) | Individuals with either static scores and a moderate or high baseline severity or those with worsening scores. |
| Integrated Baseline Mood Severity | Combined baseline mood severity across PHQ, GAD, and ASRM scores. "Low" if no more than one score is low/minimal; "Not Low" if two or more scores are not low/minimal. |
| Stable/Recovering (Integrated Measure) | Individuals with at least two static scores and a low integrated baseline mood severity, or those with at least two recovering scores. |
| Resistant/Worsening (Integrated Measure) | Individuals with at least two static scores and a moderate or high integrated baseline mood severity, or those with at least two worsening scores. |
| Frequent Combined Use of Antidepressants and Antipsychotics | Individuals with an antidepressant use frequency of ≥0.5 and an antipsychotic use frequency of ≥0.3, with thresholds chosen based on the distribution of medication use frequency, approximately 1 standard deviation above the mean. |

| | |
|---|---|
| Frequent Antidepressant Use Only | Individuals with an antidepressant use frequency of ≥0.5 and an antipsychotic use frequency of <0.3. |

### S4. Comparison of LAMP and LCGA

In comparing the methodologies employed in our study with traditional approaches, it is crucial to highlight the advantages of our proposed model. One traditional approach is the latent class growth analysis (LCGA; Jung and Wickrama, 2008), commonly used in psychiatric research to categorize participants into distinct illness courses based on the longitudinal data. However, LCGA has limitations if applied to our context. Traditionally, studies using LCGA have opted for a small number of trajectory classes, typically 3-4, which oversimplifies the complexity of mood trajectories (Birmaher et al., 2014; Frías et al., 2017; Mignogna and Goes, 2024; Weintraub et al., 2020a, 2020b). For example, considering four clinically significant depression severity levels and three trajectory trends (increasing, static, or decreasing), the PHQ presents 12 potential classes, far exceeding the typical 3-4. Additionally, the class selection in LCGA is often driven by models that minimize the Bayesian Information Criterion, and the interpretation of trajectories lacks standardization, often based on intuitive labels such as "ill with improving" or "predominantly ill." This method can lead to inconsistent classifications within the same category, especially when analyzing multiple mood scores.

The LAMP methodology, inspired by the utility of Linear Mixed Models in relating the behavior of a set of variables to a dependent variable, proves highly effective for time-dependent data (Gurka and Edwards, 2011). Mixed models have shown remarkable utility in analyzing relationships between time-dependent variables and affective symptoms in patients with Bipolar Disorder, revealing significant correlations between mood, self-assessment of mood, and socialization indicators (Dominiak et al., 2022). Furthermore, self-assessment of mood and other variables of interest have been used to predict the occurrence of affective episodes or euthymia using a logit generalized linear mixed model (Ludwig et al., 2024). The LAMP approach provides an additional layer of context beyond simple mixed models.

### S5. Additional Figures and Tables

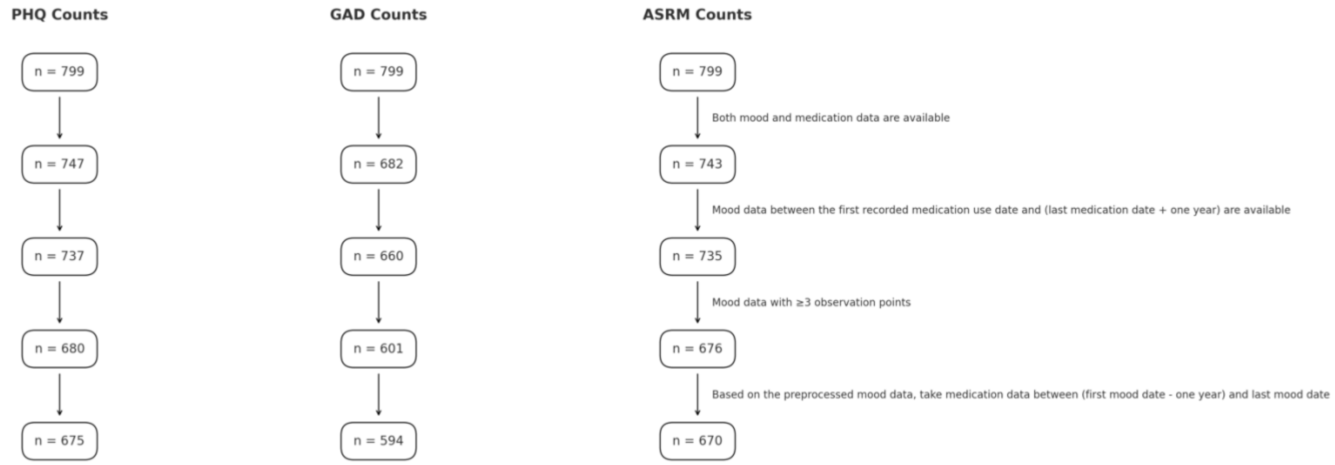


Figure S1: Flowcharts illustrating the inclusion and exclusion criteria and the resulting sample sizes for the analysis of mood data in patients with Bipolar Disorder (BP) for PHQ-9 (A), GAD-7 (B), and ASRM (C).

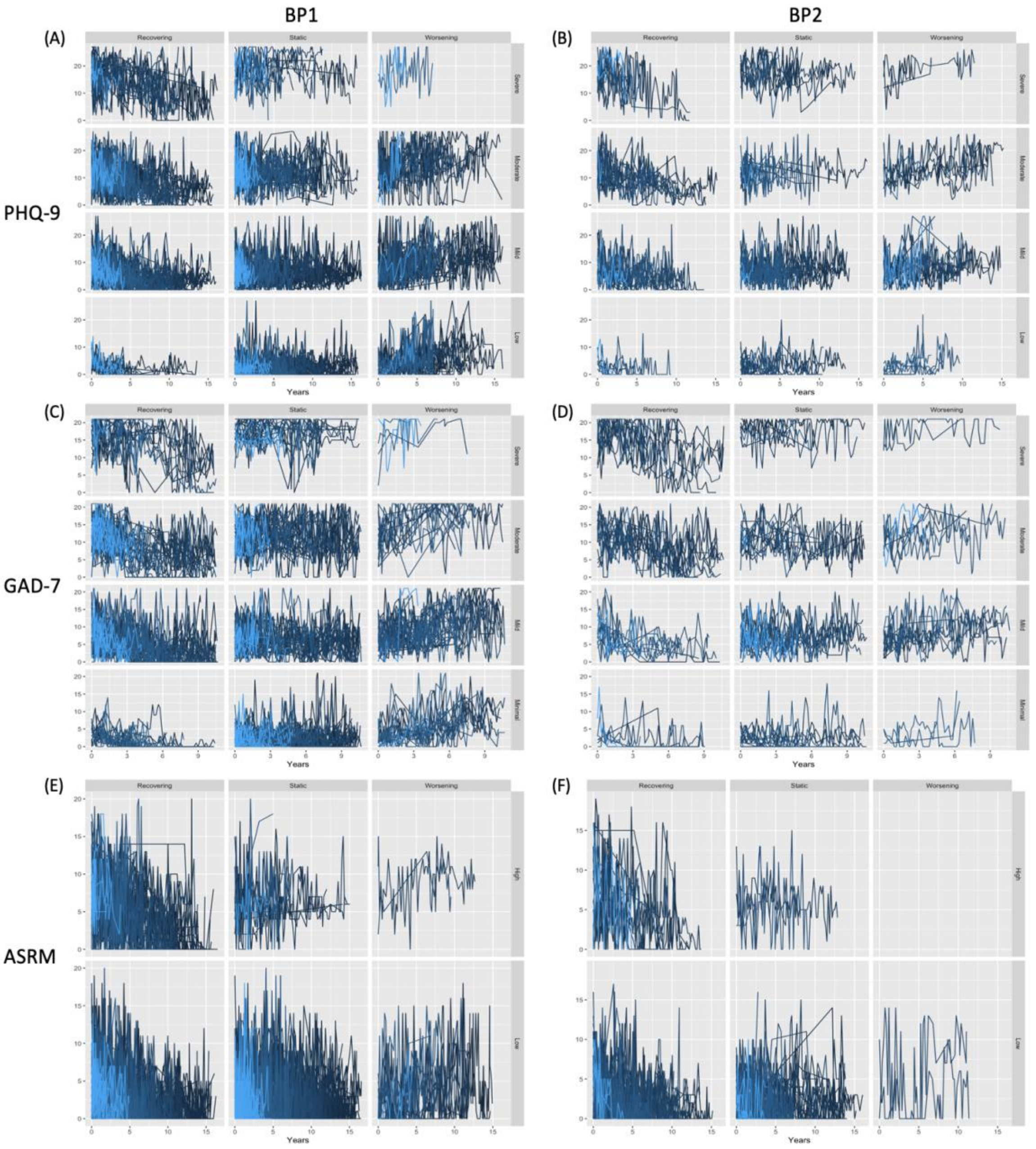


Figure S2: Visualization of Mood Trajectories. Panels A, C, and E correspond to the PHQ-9, GAD-7, and ASRM trajectories of the BP1 cohort, respectively, while Panels B, D, and F correspond to the PHQ-9, GAD-7, and ASRM trajectories of the BP2 cohort, respectively.

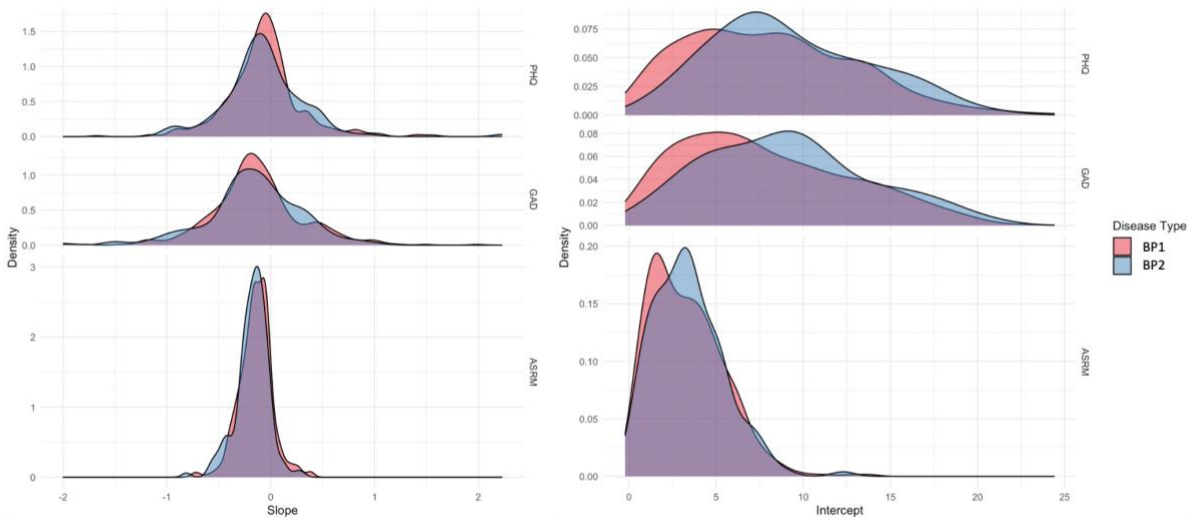


Figure S3: The density distribution of intercepts and slopes for BP1 and BP2, separated by each mood score type.

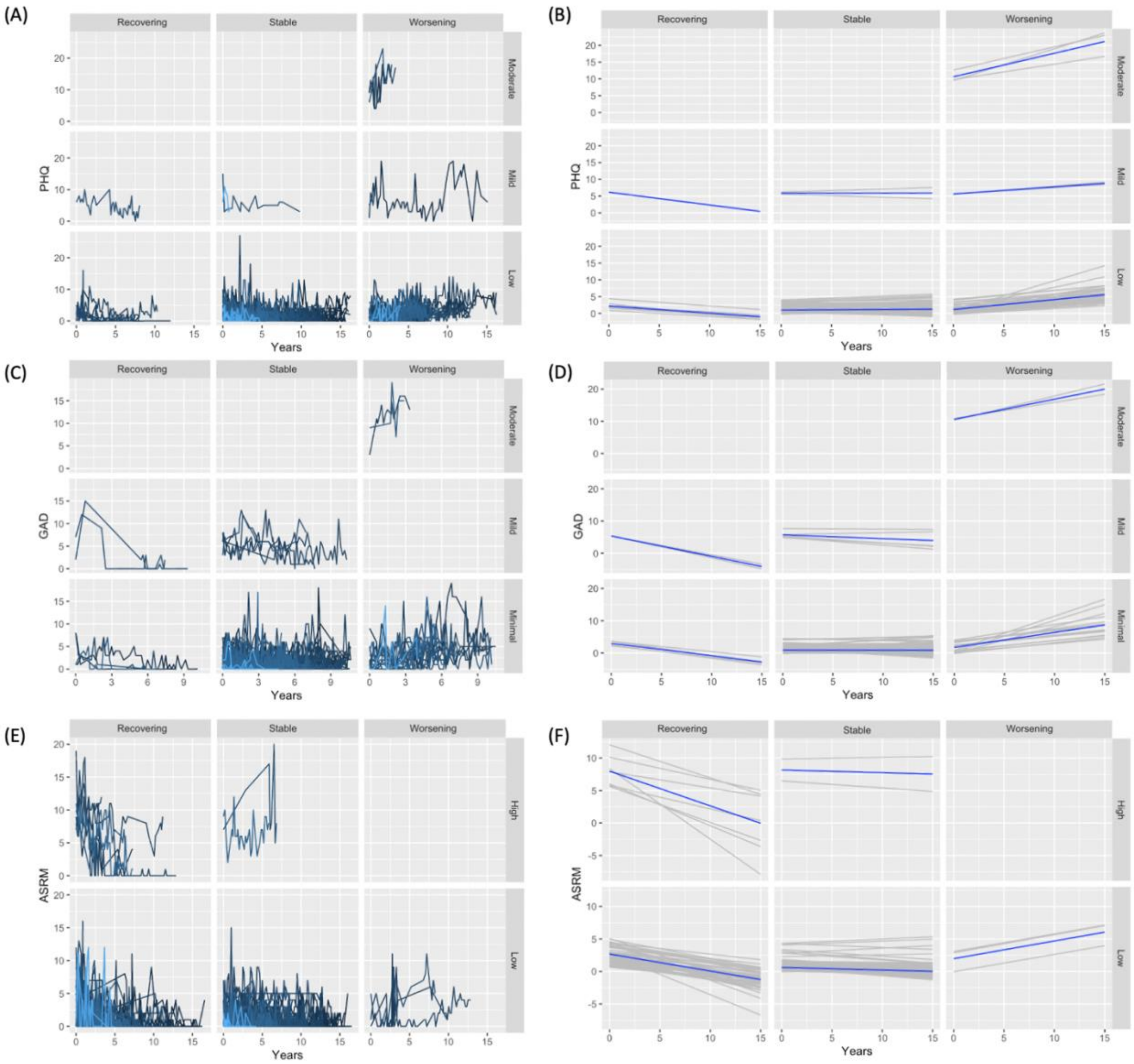


Figure S4: Visualization of Individual and Smoothed Trajectories for PHQ, GAD, and ASRM Scores in the healthy control group. Panels A, C, and E depict the raw trajectories, while Panels B, D, and F display their corresponding smoothed trends. These trajectories are categorized into subgroups based on different levels of baseline severity and symptom progression.

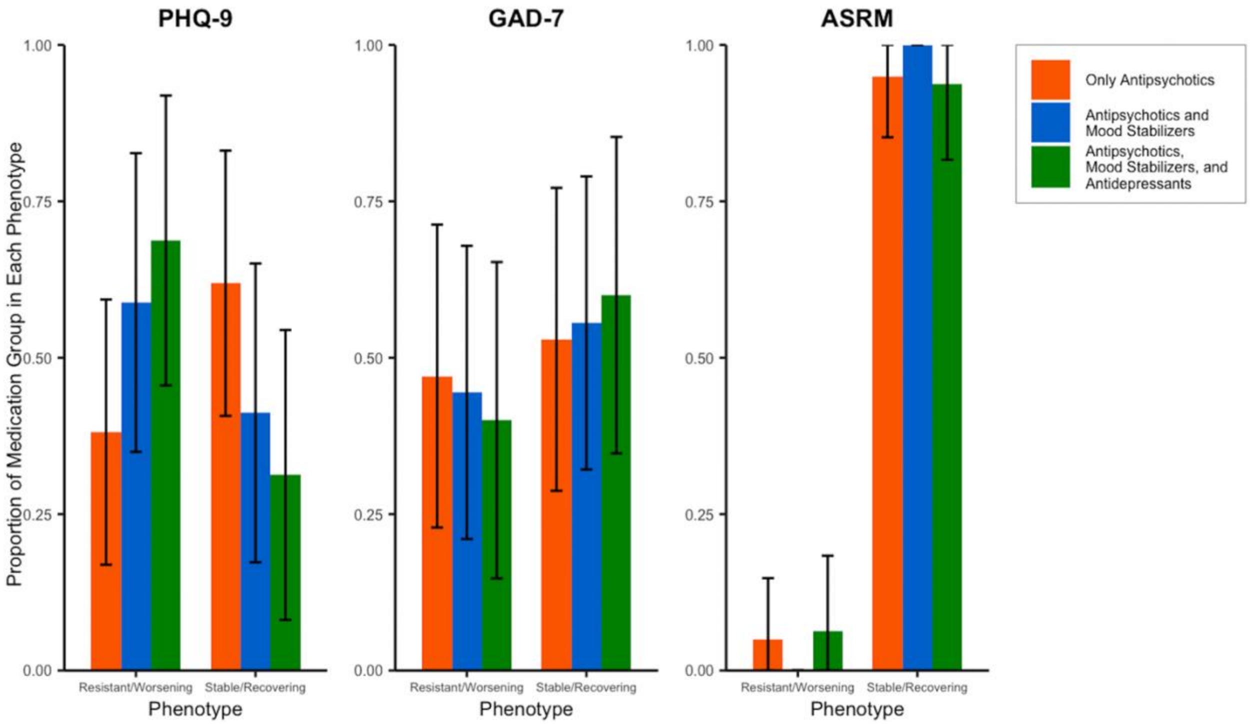


Figure S5. LAMP Phenotype Distributions within Bipolar 1. The figure shows the proportion of individuals within each phenotype (Resistant/Worsening and Stable/Recovering) across three mood measures: PHQ-9, GAD-7, and ASRM. The bars represent different medication groups: antipsychotics only (orange), antipsychotics and mood stabilizers (blue), and antipsychotics, mood stabilizers, and antidepressants (green). The error bars represent 95% confidence intervals for each group. The distribution patterns illustrate the effects of these medication groups on mood outcomes within the Bipolar 1 cohort.

| | | BP1 (N=525) | BP2 (N=150) | Healthy Control (N=208) | P-value |
|---|---|---|---|---|---|
| Demographics | **Gender** | | | | |
| | Female | 346 (65.9%) | 111 (74.0%) | 147 (70.7%) | Chi-sq, P=0.12 |
| | Male | 179 (34.1%) | 39 (26.0%) | 61 (29.3%) | |
| | **Age at Baseline** | | | | |
| | Mean (SD) | 41 (± 13) | 43 (± 15) | 40 (± 16) | ANOVA, P=0.11 |
| | **Marital Status at Baseline** | | | | |
| | Divorced/Separated/Widowed | 104 (19.8%) | 37 (24.7%) | 23 (11.1%) | Chi-sq, P=0.0076 |
| | Married | 197 (37.5%) | 60 (40.0%) | 83 (39.9%) | |
| | Never Married | 224 (42.7%) | 53 (35.3%) | 102 (49.0%) | |
| Medication | **Average Number of Concurrent Medications** | | | | |
| | Mean (SD) | 6.2 (± 3.8) | 6.1 (± 3.7) | 2.7 (± 2.4) | ANOVA, P<.001 |
| | **Antipsychotics Use Frequency** | | | | |
| | Mean (SD) | 0.13 (± 0.23) | 0.088 (± 0.18) | 0 (± 0) | ANOVA, P<.001 |
| | **Antidepressants Use Frequency** | | | | |
| | Mean (SD) | 0.20 (± 0.31) | 0.27 (± 0.32) | 0.017 (± 0.084) | ANOVA, P<.001 |
| | **Anxiolytics Use Frequency** | | | | |
| | Mean (SD) | 0.062 (± 0.17) | 0.059 (± 0.13) | 0.0090 (± 0.075) | ANOVA, P<.001 |
| | **Mood Stabilizer Use Frequency** | | | | |
| | Mean (SD) | 0.44 (± 0.41) | 0.33 (± 0.37) | 0.0054 (± 0.041) | ANOVA, P<.001 |
| Substance Use Disorders | **Alcohol Use Disorder** | | | | |
| | Yes | 224 (42.7%) | 55 (36.7%) | 1 (0.5%) | Chi-sq, P<.001 |
| | No | 301 (57.3%) | 95 (63.3%) | 207 (99.5%) | |
| | **Cannabis Use Disorder** | | | | |
| | Yes | 149 (28.4%) | 32 (21.3%) | 0 (0%) | Chi-sq, P<.001 |
| | No | 376 (71.6%) | 118 (78.7%) | 208 (100%) | |
| | **Cocaine Use Disorder** | | | | |
| | Yes | 50 (9.5%) | 12 (8.0%) | 0 (0%) | Chi-sq, P<.001 |
| | No | 475 (90.5%) | 138 (92.0%) | 208 (100%) | |
| Comorbidity | **Specific Phobia** | | | | |
| | Yes | 52 (9.9%) | 22 (14.7%) | 0 (0%) | Chi-sq, P<.001 |
| | No | 473 (90.1%) | 128 (85.3%) | 208 (100%) | |
| | **PTSD** | | | | |
| | Yes | 67 (12.8%) | 20 (13.3%) | 0 (0%) | Chi-sq, P<.001 |
| | No | 458 (87.2%) | 130 (86.7%) | 208 (100%) | |
| | **Head Injury** | | | | |
| | Yes | 118 (22.5%) | 25 (16.7%) | 12 (5.8%) | Chi-sq, P<.001 |
| | No | 309 (58.9%) | 92 (61.3%) | 180 (86.5%) | |
| | Missing | 98 (18.7%) | 33 (22.0%) | 16 (7.7%) | |
| | **Migraine** | | | | |
| | Yes | 132 (25.1%) | 51 (34.0%) | 25 (12.0%) | Chi-sq, P<.001 |
| | No | 297 (56.6%) | 67 (44.7%) | 166 (79.8%) | |
| | Missing | 96 (18.3%) | 32 (21.3%) | 17 (8.2%) | |
| | **Overweight** | | | | |
| | Yes | 218 (41.5%) | 56 (37.3%) | 31 (14.9%) | Chi-sq, P<.001 |
| | No | 211 (40.2%) | 61 (40.7%) | 160 (76.9%) | |
| | Missing | 96 (18.3%) | 33 (22.0%) | 17 (8.2%) | |

Table S1: Summary statistics for the BP1, BP2, and healthy control cohorts. For continuous covariates, means and standard deviations are provided. For categorical variables, sample sizes and corresponding proportions are provided.

| Mood Measure | Baseline Severity | Progression | BP1 | | | BP2 | | |
|---|---|---|---|---|---|---|---|---|
| | | | Sample Size | Average Intercept | Average Slope | Sample Size | Average Intercept | Average Slope |
| PHQ-9 | Low | Recovering | 13 | 3.56 | -0.29 | 5 | 3.18 | -0.27 |
| | Low | Static | 105 | 2.31 | 0.01 | 12 | 3.38 | -0.04 |
| | Low | Worsening | 25 | 3.08 | 0.52 | 6 | 2.45 | 0.39 |
| | Mild | Recovering | 64 | 7.22 | -0.39 | 20 | 7.34 | -0.31 |
| | Mild | Static | 93 | 6.76 | -0.03 | 29 | 7.19 | -0.02 |
| | Mild | Worsening | 32 | 7.12 | 0.49 | 16 | 6.73 | 0.52 |
| | Moderate | Recovering | 62 | 12.01 | -0.44 | 18 | 12.45 | -0.47 |
| | Moderate | Static | 59 | 11.54 | -0.04 | 14 | 11.63 | -0.02 |
| | Moderate | Worsening | 21 | 12.29 | 0.57 | 7 | 11.26 | 0.36 |
| | Severe | Recovering | 33 | 17.31 | -0.49 | 12 | 17.02 | -0.55 |
| | Severe | Static | 16 | 17.74 | -0.03 | 8 | 17.37 | -0.06 |
| | Severe | Worsening | 2 | 16.47 | 0.56 | 3 | 15.74 | 0.45 |
| GAD-7 | Minimal | Recovering | 20 | 3.33 | -0.39 | 7 | 3.03 | -0.38 |
| | Minimal | Static | 100 | 2.29 | -0.06 | 16 | 2.60 | -0.09 |
| | Minimal | Worsening | 17 | 3.28 | 0.60 | 3 | 3.13 | 0.41 |
| | Mild | Recovering | 64 | 6.91 | -0.53 | 14 | 7.47 | -0.51 |
| | Mild | Static | 65 | 6.63 | -0.08 | 25 | 7.08 | -0.04 |
| | Mild | Worsening | 33 | 6.79 | 0.64 | 12 | 6.79 | 0.42 |
| | Moderate | Recovering | 48 | 12.00 | -0.63 | 17 | 11.63 | -0.64 |
| | Moderate | Static | 45 | 11.55 | -0.06 | 16 | 10.99 | -0.08 |
| | Moderate | Worsening | 13 | 12.41 | 0.61 | 7 | 11.12 | 0.55 |
| | Severe | Recovering | 24 | 17.18 | -0.67 | 13 | 17.10 | -0.82 |
| | Severe | Static | 22 | 16.74 | -0.08 | 7 | 17.14 | -0.02 |
| | Severe | Worsening | 4 | 15.84 | 0.51 | 2 | 16.05 | 0.53 |
| ASRM | Low | Recovering | 170 | 3.60 | -0.24 | 63 | 3.52 | -0.25 |
| | Low | Static | 251 | 1.99 | -0.06 | 66 | 2.25 | -0.06 |
| | Low | Worsening | 16 | 2.22 | 0.27 | 2 | 2.92 | 0.27 |
| | High | Recovering | 68 | 7.01 | -0.36 | 15 | 7.42 | -0.42 |
| | High | Static | 13 | 6.41 | -0.06 | 4 | 6.05 | -0.11 |
| | High | Worsening | 2 | 6.98 | 0.22 | 0 | | |

Table S2: Summary statistics for all classified categories. For each mood score, the classified category is characterized by baseline severity and level of progression. For each category, the table provides the sample size, average estimated intercept, and average estimated slope of the linear trend for each subgroup, separately for BP1 and BP2.

| | PHQ-9 | | GAD-7 | | ASRM | | Integrated | |
|---|---|---|---|---|---|---|---|---|
| | Stable/Recovering (N=277) | Resistant/Worsening (N=248) | Stable/Recovering (N=256) | Resistant/Worsening (N=199) | Stable/Recovering (N=489) | Resistant/Worsening (N=31) | Stable/Recovering (N=238) | Resistant/Worsening (N=177) |
| **Gender** | | | | | | | | |
| Female | 183 (66.1%) | 163 (65.7%) | 170 (66.4%) | 138 (69.3%) | 327 (66.9%) | 18 (58.1%) | 163 (68.5%) | 118 (66.7%) |
| Male | 94 (33.9%) | 85 (34.3%) | 86 (33.6%) | 61 (30.7%) | 162 (33.1%) | 13 (41.9%) | 75 (31.5%) | 59 (33.3%) |
| **Age at Baseline** | | | | | | | | |
| Mean (SD) | 42 (± 14) | 40 (± 13) | 44 (± 14) | 41 (± 12) | 41 (± 13) | 45 (± 13) | 42 (± 14) | 39 (± 13) |
| **Marital Status at Baseline** | | | | | | | | |
| Divorced/Separated/Widowed | 55 (19.9%) | 49 (19.8%) | 49 (19.1%) | 38 (19.1%) | 101 (20.7%) | 3 (9.7%) | 46 (19.3%) | 35 (19.8%) |
| Married | 116 (41.9%) | 81 (32.7%) | 107 (41.8%) | 69 (34.7%) | 178 (36.4%) | 17 (54.8%) | 99 (41.6%) | 61 (34.5%) |
| Never Married | 106 (38.3%) | 118 (47.6%) | 100 (39.1%) | 92 (46.2%) | 210 (42.9%) | 11 (35.5%) | 93 (39.1%) | 81 (45.8%) |
| **Average Number of Concurrent Medications** | | | | | | | | |
| Mean (SD) | 5.8 (± 3.4) | 6.7 (± 4.1) | 6.3 (± 3.9) | 6.9 (± 4.0) | 6.2 (± 3.7) | 7.1 (± 4.4) | 6.0 (± 3.7) | 6.9 (± 3.9) |
| **Antipsychotics Use Frequency** | | | | | | | | |
| Mean (SD) | 0.11 (± 0.20) | 0.14 (± 0.26) | 0.12 (± 0.22) | 0.16 (± 0.27) | 0.12 (± 0.23) | 0.12 (± 0.22) | 0.12 (± 0.21) | 0.16 (± 0.27) |
| **Antidepressants Use Frequency** | | | | | | | | |
| Mean (SD) | 0.21 (± 0.31) | 0.19 (± 0.31) | 0.22 (± 0.33) | 0.20 (± 0.29) | 0.20 (± 0.31) | 0.19 (± 0.25) | 0.21 (± 0.31) | 0.20 (± 0.30) |
| **Anxiolytics Use Frequency** | | | | | | | | |
| Mean (SD) | 0.054 (± 0.16) | 0.071 (± 0.18) | 0.071 (± 0.19) | 0.069 (± 0.17) | 0.063 (± 0.17) | 0.061 (± 0.16) | 0.057 (± 0.17) | 0.083 (± 0.19) |
| **Mood Stabilizer Use Frequency** | | | | | | | | |
| Mean (SD) | 0.44 (± 0.41) | 0.44 (± 0.40) | 0.49 (± 0.42) | 0.43 (± 0.40) | 0.44 (± 0.41) | 0.43 (± 0.39) | 0.48 (± 0.41) | 0.47 (± 0.39) |
| **Alcohol Use Disorder** | | | | | | | | |
| Yes | 101 (36.5%) | 123 (49.6%) | 84 (32.8%) | 110 (55.3%) | 204 (41.7%) | 17 (54.8%) | 83 (34.9%) | 93 (52.5%) |
| No | 176 (63.5%) | 125 (50.4%) | 172 (67.2%) | 89 (44.7%) | 285 (58.3%) | 14 (45.2%) | 155 (65.1%) | 84 (47.5%) |
| **Cannabis Use Disorder** | | | | | | | | |
| Yes | 79 (28.5%) | 70 (28.2%) | 63 (24.6%) | 68 (34.2%) | 135 (27.6%) | 10 (32.3%) | 63 (26.5%) | 56 (31.6%) |
| No | 198 (71.5%) | 178 (71.8%) | 193 (75.4%) | 131 (65.8%) | 354 (72.4%) | 21 (67.7%) | 175 (73.5%) | 121 (68.4%) |
| **Cocaine Use Disorder** | | | | | | | | |
| Yes | 26 (9.4%) | 24 (9.7%) | 18 (7.0%) | 26 (13.1%) | 46 (9.4%) | 3 (9.7%) | 20 (8.4%) | 19 (10.7%) |
| No | 251 (90.6%) | 224 (90.3%) | 238 (93.0%) | 173 (86.9%) | 443 (90.6%) | 28 (90.3%) | 218 (91.6%) | 158 (89.3%) |
| **Specific Phobia** | | | | | | | | |
| Yes | 23 (8.3%) | 29 (11.7%) | 23 (9.0%) | 20 (10.1%) | 51 (10.4%) | 1 (3.2%) | 20 (8.4%) | 17 (9.6%) |
| No | 254 (91.7%) | 219 (88.3%) | 233 (91.0%) | 179 (89.9%) | 438 (89.6%) | 30 (96.8%) | 218 (91.6%) | 160 (90.4%) |
| **PTSD** | | | | | | | | |
| Yes | 29 (10.5%) | 38 (15.3%) | 30 (11.7%) | 33 (16.6%) | 63 (12.9%) | 3 (9.7%) | 28 (11.8%) | 28 (15.8%) |
| No | 248 (89.5%) | 210 (84.7%) | 226 (88.3%) | 166 (83.4%) | 426 (87.1%) | 28 (90.3%) | 210 (88.2%) | 149 (84.2%) |
| **Head Injury** | | | | | | | | |
| Yes | 62 (22.4%) | 56 (22.6%) | 60 (23.4%) | 36 (18.1%) | 108 (22.1%) | 8 (25.8%) | 54 (22.7%) | 32 (18.1%) |
| No | 179 (64.6%) | 130 (52.4%) | 148 (57.8%) | 117 (58.8%) | 293 (59.9%) | 14 (45.2%) | 145 (60.9%) | 96 (54.2%) |
| Missing | 36 (13.0%) | 62 (25.0%) | 48 (18.8%) | 46 (23.1%) | 88 (18.0%) | 9 (29.0%) | 39 (16.4%) | 49 (27.7%) |
| **Migraine** | | | | | | | | |
| Yes | 66 (23.8%) | 66 (26.6%) | 60 (23.4%) | 51 (25.6%) | 126 (25.8%) | 6 (19.4%) | 59 (24.8%) | 47 (26.6%) |
| No | 173 (62.5%) | 124 (50.0%) | 147 (57.4%) | 105 (52.8%) | 277 (56.6%) | 16 (51.6%) | 139 (58.4%) | 84 (47.5%) |
| Missing | 38 (13.7%) | 58 (23.4%) | 49 (19.1%) | 43 (21.6%) | 86 (17.6%) | 9 (29.0%) | 40 (16.8%) | 46 (26.0%) |
| **Overweight** | | | | | | | | |
| Yes | 115 (41.5%) | 103 (41.5%) | 102 (39.8%) | 84 (42.2%) | 202 (41.3%) | 14 (45.2%) | 96 (40.3%) | 69 (39.0%) |
| No | 124 (44.8%) | 87 (35.1%) | 105 (41.0%) | 72 (36.2%) | 201 (41.1%) | 8 (25.8%) | 102 (42.9%) | 62 (35.0%) |
| Missing | 38 (13.7%) | 58 (23.4%) | 49 (19.1%) | 43 (21.6%) | 86 (17.6%) | 9 (29.0%) | 40 (16.8%) | 46 (26.0%) |

Table S3: Summary statistics for the BP1 cohort, presenting separate comparisons between the "Stable/Recovering" and "Resistant/Worsening" phenotype groups across individual mood scores and their integrated measure. For continuous covariates, means and standard deviations are provided. For categorical variables, sample sizes and corresponding proportions are provided.

| | PHQ-9 | | GAD-7 | | ASRM | | Integrated | |
|---|---|---|---|---|---|---|---|---|
| | Stable/Recovering (N=67) | Resistant/Worsening (N=83) | Stable/Recovering (N=67) | Resistant/Worsening (N=72) | Stable/Recovering (N=144) | Resistant/Worsening (N=6) | Stable/Recovering (N=65) | Resistant/Worsening (N=49) |
| **Gender** | | | | | | | | |
| Female | 53 (79.1%) | 58 (69.9%) | 50 (74.6%) | 54 (75.0%) | 107 (74.3%) | 4 (66.7%) | 48 (73.8%) | 37 (75.5%) |
| Male | 14 (20.9%) | 25 (30.1%) | 17 (25.4%) | 18 (25.0%) | 37 (25.7%) | 2 (33.3%) | 17 (26.2%) | 12 (24.5%) |
| **Age at Baseline** | | | | | | | | |
| Mean (SD) | 44 (± 15) | 42 (± 15) | 48 (± 15) | 42 (± 15) | 43 (± 15) | 38 (± 14) | 45 (± 16) | 40 (± 13) |
| **Marital Status at Baseline** | | | | | | | | |
| Divorced/Separated/Widowed | 17 (25.4%) | 20 (24.1%) | 20 (29.9%) | 16 (22.2%) | 35 (24.3%) | 3 (50.0%) | 18 (27.7%) | 9 (18.4%) |
| Married | 28 (41.8%) | 32 (38.6%) | 29 (43.3%) | 26 (36.1%) | 59 (41.0%) | 1 (16.7%) | 26 (40.0%) | 22 (44.9%) |
| Never Married | 22 (32.8%) | 31 (37.3%) | 18 (26.9%) | 30 (41.7%) | 50 (34.7%) | 2 (33.3%) | 21 (32.3%) | 18 (36.7%) |
| **Average Number of Concurrent Medications** | | | | | | | | |
| Mean (SD) | 5.5 (± 2.8) | 6.5 (± 4.3) | 6.1 (± 3.6) | 6.7 (± 4.3) | 6.2 (± 3.8) | 4.8 (± 1.8) | 5.9 (± 3.5) | 6.3 (± 3.5) |
| **Antipsychotics Use Frequency** | | | | | | | | |
| Mean (SD) | 0.12 (± 0.22) | 0.066 (± 0.14) | 0.13 (± 0.23) | 0.076 (± 0.14) | 0.088 (± 0.18) | 0.069 (± 0.17) | 0.14 (± 0.24) | 0.056 (± 0.12) |
| **Antidepressants Use Frequency** | | | | | | | | |
| Mean (SD) | 0.29 (± 0.35) | 0.26 (± 0.31) | 0.32 (± 0.35) | 0.27 (± 0.33) | 0.26 (± 0.32) | 0.47 (± 0.34) | 0.29 (± 0.34) | 0.31 (± 0.34) |
| **Anxiolytics Use Frequency** | | | | | | | | |
| Mean (SD) | 0.040 (± 0.12) | 0.073 (± 0.14) | 0.075 (± 0.18) | 0.066 (± 0.14) | 0.057 (± 0.13) | 0.083 (± 0.14) | 0.049 (± 0.12) | 0.084 (± 0.16) |
| **Mood Stabilizer Use Frequency** | | | | | | | | |
| Mean (SD) | 0.32 (± 0.34) | 0.33 (± 0.39) | 0.40 (± 0.39) | 0.31 (± 0.36) | 0.32 (± 0.36) | 0.42 (± 0.47) | 0.38 (± 0.36) | 0.34 (± 0.38) |
| **Alcohol Use Disorder** | | | | | | | | |
| Yes | 29 (43.3%) | 26 (31.3%) | 20 (29.9%) | 27 (37.5%) | 52 (36.1%) | 2 (33.3%) | 21 (32.3%) | 17 (34.7%) |
| No | 38 (56.7%) | 57 (68.7%) | 47 (70.1%) | 45 (62.5%) | 92 (63.9%) | 4 (66.7%) | 44 (67.7%) | 32 (65.3%) |
| **Cannabis Use Disorder** | | | | | | | | |
| Yes | 15 (22.4%) | 17 (20.5%) | 14 (20.9%) | 13 (18.1%) | 30 (20.8%) | 1 (16.7%) | 12 (18.5%) | 7 (14.3%) |
| No | 52 (77.6%) | 66 (79.5%) | 53 (79.1%) | 59 (81.9%) | 114 (79.2%) | 5 (83.3%) | 53 (81.5%) | 42 (85.7%) |
| **Cocaine Use Disorder** | | | | | | | | |
| Yes | 6 (9.0%) | 6 (7.2%) | 6 (9.0%) | 6 (8.3%) | 10 (6.9%) | 1 (16.7%) | 5 (7.7%) | 4 (8.2%) |
| No | 61 (91.0%) | 77 (92.8%) | 61 (91.0%) | 66 (91.7%) | 134 (93.1%) | 5 (83.3%) | 60 (92.3%) | 45 (91.8%) |
| **Specific Phobia** | | | | | | | | |
| Yes | 8 (11.9%) | 14 (16.9%) | 9 (13.4%) | 12 (16.7%) | 21 (14.6%) | 1 (16.7%) | 9 (13.8%) | 9 (18.4%) |
| No | 59 (88.1%) | 69 (83.1%) | 58 (86.6%) | 60 (83.3%) | 123 (85.4%) | 5 (83.3%) | 56 (86.2%) | 40 (81.6%) |
| **PTSD** | | | | | | | | |
| Yes | 9 (13.4%) | 11 (13.3%) | 9 (13.4%) | 11 (15.3%) | 19 (13.2%) | 0 (0%) | 8 (12.3%) | 7 (14.3%) |
| No | 58 (86.6%) | 72 (86.7%) | 58 (86.6%) | 61 (84.7%) | 125 (86.8%) | 6 (100%) | 57 (87.7%) | 42 (85.7%) |
| **Head Injury** | | | | | | | | |
| Yes | 10 (14.9%) | 15 (18.1%) | 7 (10.4%) | 15 (20.8%) | 24 (16.7%) | 1 (16.7%) | 9 (13.8%) | 9 (18.4%) |
| No | 43 (64.2%) | 49 (59.0%) | 44 (65.7%) | 41 (56.9%) | 88 (61.1%) | 4 (66.7%) | 40 (61.5%) | 30 (61.2%) |
| Missing | 14 (20.9%) | 19 (22.9%) | 16 (23.9%) | 16 (22.2%) | 32 (22.2%) | 1 (16.7%) | 16 (24.6%) | 10 (20.4%) |
| **Migraine** | | | | | | | | |
| Yes | 19 (28.4%) | 32 (38.6%) | 22 (32.8%) | 25 (34.7%) | 47 (32.6%) | 3 (50.0%) | 19 (29.2%) | 20 (40.8%) |
| No | 34 (50.7%) | 33 (39.8%) | 29 (43.3%) | 32 (44.4%) | 66 (45.8%) | 2 (33.3%) | 30 (46.2%) | 20 (40.8%) |
| Missing | 14 (20.9%) | 18 (21.7%) | 16 (23.9%) | 15 (20.8%) | 31 (21.5%) | 1 (16.7%) | 16 (24.6%) | 9 (18.4%) |
| **Overweight** | | | | | | | | |
| Yes | 22 (32.8%) | 34 (41.0%) | 25 (37.3%) | 28 (38.9%) | 53 (36.8%) | 3 (50.0%) | 20 (30.8%) | 23 (46.9%) |
| No | 31 (46.3%) | 30 (36.1%) | 25 (37.3%) | 29 (40.3%) | 59 (41.0%) | 2 (33.3%) | 28 (43.1%) | 17 (34.7%) |
| Missing | 14 (20.9%) | 19 (22.9%) | 17 (25.4%) | 15 (20.8%) | 32 (22.2%) | 1 (16.7%) | 17 (26.2%) | 9 (18.4%) |

Table S4: Summary statistics for the BP2 cohort, presenting separate comparisons between the "Stable/Recovering" and "Resistant/Worsening" phenotype groups across individual mood scores and their integrated measure. For continuous covariates, means and standard deviations are provided. For categorical variables, sample sizes and corresponding proportions are provided.

References (Supplementary Materials)


Birmaher, B., Gill, M.K., Axelson, D.A., Goldstein, B.I., Goldstein, T.R., Yu, H., Liao, F., Iyengar, S., Diler, R.S., Strober, M., Hower, H., Yen, S., Hunt, J., Merranko, J.A., Ryan, N.D., Keller, M.B., 2014. Longitudinal Trajectories and Associated Baseline Predictors in Youths With Bipolar Spectrum Disorders. Am. J. Psychiatry 171, 990–999. https://doi.org/10.1176/appi.ajp.2014.13121577

Dominiak, M., Kaczmarek-Majer, K., Antosik-Wójcińska, A.Z., Opara, K.R., Olwert, A., Radziszewska, W., Hryniewicz, O., Święcicki, Ł., Wojnar, M., Mierzejewski, P., 2022. Behavioral and self-reported data collected from smartphones for the assessment of depressive and manic symptoms in patients with bipolar disorder: Prospective observational study. J. Med. Internet Res. 24, e28647. https://doi.org/10.2196/28647

Frías, Á., Dickstein, D.P., Merranko, J., Gill, M.K., Goldstein, T.R., Goldstein, B.I., Hower, H., Yen, S., Hafeman, D.M., Liao, F., Diler, R., Axelson, D., Strober, M., Hunt, J.I., Ryan, N.D., Keller, M.B., Birmaher, B., 2017. Longitudinal cognitive trajectories and associated clinical variables in youth with bipolar disorder. Bipolar Disord. 19, 273–284. https://doi.org/10.1111/bdi.12510

Gurka, M.J., Edwards, L.J., 2011. Mixed Models, in: Essential Statistical Methods for Medical Statistics. Elsevier, pp. 146–173. https://doi.org/10.1016/b978-0-444-53737-9.50008-6

Jung, T., Wickrama, K.A.S., 2008. An Introduction to Latent Class Growth Analysis and Growth Mixture Modeling. Soc. Personal. Psychol. Compass 2, 302–317. https://doi.org/10.1111/j.1751-9004.2007.00054.x

Ludwig, V.M., Reinhard, I., Mühlbauer, E., Hill, H., Severus, W.E., Bauer, M., Ritter, P., Ebner-Priemer, U.W., 2024. Limited evidence of autocorrelation signaling upcoming affective episodes: a 12-month e-diary study in patients with bipolar disorder. Psychol. Med. 54, 1844–1852. https://doi.org/10.1017/S0033291723003811

Mignogna, K.M., Goes, F.S., 2024. Characterizing the longitudinal course of symptoms and functioning in bipolar disorder. Psychol. Med. 54, 79–89. https://doi.org/10.1017/S0033291722001489

Weintraub, M.J., Schneck, C.D., Axelson, D.A., Birmaher, B., Kowatch, R.A., Miklowitz, D.J., 2020a. Classifying Mood Symptom Trajectories in Adolescents With Bipolar Disorder. J. Am. Acad. Child Adolesc. Psychiatry 59, 381–390. https://doi.org/10.1016/j.jaac.2019.04.028

Weintraub, M.J., Schneck, C.D., Walshaw, P.D., Chang, K.D., Sullivan, A.E., Singh, M.K., Miklowitz, D.J., 2020b. Longitudinal trajectories of mood symptoms and global functioning in youth at high risk for bipolar disorder. J. Affect. Disord. 277, 394–401. https://doi.org/10.1016/j.jad.2020.08.018
